# Novel transposon Tn*8026* contributes to the global spread of transmissible linezolid resistance in *Enterococcus* via a linear plasmid

**DOI:** 10.64898/2026.03.04.26347163

**Authors:** Michael B Hall, Yiyang Xue, Tricia S.E Lee, Ella Herring, Jocelyn Hume, Ryan R Wick, Timothy Kidd, Naomi Runnegar, Patrick N A Harris, Bianca Graves, Leah W Roberts

## Abstract

Linezolid is a critical last-resort antimicrobial for multidrug-resistant *Enterococcus faecium*, particularly against vancomycin-resistant lineages where therapeutic options are severely limited. While resistance has historically arisen through *de novo* chromosomal mutations, the global emergence of transferable resistance mechanisms threatens to render more infections untreatable. Here, we characterise a recent (2023–2024) hospital-associated outbreak of linezolid-resistant *E. faecium* in Queensland, Australia. Although the cohort comprised a variety of sequence types, the outbreak was primarily driven by the clonal expansion of an ST80 lineage carrying the plasmid-borne *poxtA* gene.

Standard short-read genomic surveillance failed to resolve the genetic context of the resistance determinant. However, long-read sequencing revealed that *poxtA* was carried within a novel transposon, Tn*8026*, situated on a linear plasmid. Structural analysis defined Tn*8026* as a unique element flanked by IS*1678* and the novel insertion sequence IS*Efa26*. Furthermore, we identified an instance of Tn*8026* integration into the chromosome, providing functional evidence of its mobility and capacity for stabilisation within the genome.

Global genomic screening demonstrated that Tn*8026* significantly predates the local outbreak, identified in a historical Norwegian isolate from 2012, indicating a long-standing yet unrecognised global reservoir. Phylogenomic analysis provided strong evidence that the linear plasmid was imported from the Indian subcontinent, initiating a chain of silent dissemination in eastern Australia where the lineage circulated undetected prior to clinical recognition. Crucially, we also confirmed the presence of the linear plasmid in *Enterococcus gallinarum*, demonstrating its capacity to mobilise transmissible linezolid resistance across enterococcal species boundaries. These findings emphasise the need for detailed long-read-based surveillance of mobile genetic elements, with a particular focus on identifying linear plasmids that are often overlooked.

## Introduction

*Enterococcus faecium* has evolved from a benign gastrointestinal commensal into a leading cause of multidrug-resistant (MDR) healthcare-associated infections globally [2]. The success of this pathogen is driven by its extraordinary genomic plasticity, which facilitates the acquisition of mobile genetic elements (MGEs) encoding resistance to key antimicrobial agents [3]. Of particular concern is the global dissemination of vancomycin-resistant *E. faecium* (VREfm). In Australia, VREfm is now endemic, with surveillance data indicating some of the highest prevalence rates in the world [4,5]. Consequently, clinical management relies heavily on a diminishing number of last-resort antibiotics, principally daptomycin and the oxazolidinone linezolid [6].

The increasing reliance on linezolid has inevitably selected for resistance. While linezolid-resistant *E. faecium* (LREfm) remains comparatively rare in Australia, national reports increased 2.3-fold in 2024 (*n*=118) compared to 2023 (*n*=51), while the mechanisms driving this resistance are evolving [5,7]. Historically, linezolid resistance was primarily attributed to chromosomal point mutations in the 23S rRNA gene (e.g., G2576T), typically arising *de novo* under prolonged therapeutic pressure [8,9]. However, the epidemiology of LRE is rapidly shifting toward transferable resistance mechanisms. The emergence of plasmid-borne linezolid resistance genes such as *cfr*, *optrA*, and *poxtA* represents a critical public health threat, as these determinants can facilitate the horizontal dissemination of resistance across diverse genetic lineages and species, potentially leading to outbreaks of untreatable infections [6,8,10,11].

Understanding the mobile vehicles driving this dissemination is essential for infection control, yet standard genomic surveillance often lacks the resolution to fully characterise them. Recent studies have highlighted the role of novel linear plasmids in the spread of vancomycin and linezolid resistance in *E. faecium* [7,11–14]. These linear replicons appear to be highly conserved and stable in *Enterococcus*, yet they are frequently fragmented or misassembled [13,15]. Therefore, the contribution of linear plasmids to the stealth propagation of multidrug resistance remains underappreciated, and the transmission dynamics of these elements between international reservoirs and local hospital networks are poorly understood.

Here, we combine short- and long-read genomics to characterise a nosocomial outbreak of LREfm in a tertiary referral hospital in Queensland, Australia. We describe the emergence of a sequence type 80 (ST80) lineage carrying a novel *poxtA*-harbouring transposon, Tn*8026*, on a conserved linear plasmid. Beyond the local outbreak, we use global genomic datasets to reconstruct the intercontinental transmission history of this lineage, revealing a hidden dissemination pathway from the Indian subcontinent to eastern Australia.

## Methods

### Queensland *Enterococcus* dataset

Pathology Queensland is a statewide public pathology service supporting all Queensland public hospitals. All *Enterococcus* sp. isolates from clinical and screening samples with confirmed linezolid resistance on phenotypic antimicrobial susceptibility testing from 2023 – 2025 were included. Culture-based species identification was performed by MALDI-TOF using the Vitek-MS system and Vitek Knowledge Base Library (bioMérieux, France). Initial susceptibility testing was performed on the VITEK2 (bioMérieux, France) automated system using the P643 susceptibility panel. Resistance to linezolid was confirmed using Etest (bioMerieux, France) using EUCAST breakpoints (v11.0; resistance > 4 mg/L).

### Isolate sequencing

Short-read sequencing was performed using the Illumina MiniSeq platform. Following genomic screening with SnapperRocks (https://github.com/FordeGenomics/SnapperRocks) and LRE-Finder [16], 29 linezolid-resistant isolates were selected for long-read sequencing. High-molecular-weight DNA was extracted using the QIAamp® PowerFecal® Pro DNA Kit (QIAGEN) and sequenced on the PromethION 2 Solo (Oxford Nanopore Technologies). Raw data were basecalled with Dorado (v1.1.1) using the super-accuracy model. See Supplementary Section S1 for full details.

### Genome assembly

Genomes were assembled using Autocycler (v0.5.1; [17]), with long reads shorter than 1000 base pairs removed. All linear plasmids required manual curation in Bandage (v0.9.0; [18]) to resolve divergent paths, followed by hybrid reassembly with Unicycler (v0.5.1; [19]) to maximise terminal sequence recovery. Circular replicons were reoriented using dnaapler (v1.3.0; [20]). All assemblies were polished sequentially with long reads using Medaka (v2.1.0) and short reads using Polypolish (v0.6.0; [21]) and Pypolca (v0.6.0; [22]). See Supplementary Section S2 for full details.

### Quality control, annotation and typing

Quality of the assemblies was assessed using CheckM2 (v1.1.0; [23]). Sequence types (STs) were assigned using the mlst tool (v2.25.0) with the PubMLST database [24]. Assemblies were annotated with Bakta (v1.11.3; database v6; [25]), followed by annotation of the hypothetical proteins with Baktfold (v0.0.3; [26]).

### Global genomic dataset curation

To contextualise the study isolates, we compiled a background dataset from the Queensland Genomics Health Alliance (QGHA; [27]), the Victorian Microbiological Diagnostic Unit Public Health Laboratory (MDU PHL; [7]), the ‘AllTheBacteria’ (ATB; [28]) archive, and the National Center for Biotechnology Information’s (NCBI) GenBank [29,30]. To prevent phylogenetic overrepresentation of wild-type strains from the global databases (ATB and GenBank), we applied a targeted sampling strategy. We extracted all *Enterococcus faecium* genomes sharing ≥99.8% average nucleotide identity (ANI) with our study cohort using skani (v0.3.1; [31]). These isolates were then screened for daptomycin and linezolid resistance using AMRFinderPlus (v4.0.23; database version 2025-07-16.1; [32]), leveraging pre-computed results for ATB. Finally, susceptible isolates within the ATB and GenBank cohorts were randomly subsampled to exactly match their respective resistant counts, yielding 394 ATB and 682 GenBank genomes (see Supplementary Section S3).

### Characterisation of the *poxtA* genetic context

To characterise the genetic context of linezolid resistance, we extracted the *poxtA* coding sequence and 5,000 bp of flanking regions from annotated assemblies. Transposon boundaries were defined by identifying terminal inverted repeats (IRs): downstream IS*1678* IRs were referenced against ISfinder [33], while the novel upstream IS*1380*-family element (IS*Efa26*) IRs were determined via BLASTn (v2.15.0; [34]) self-alignment of the flanking regions.

The transposon’s global distribution was assessed by screening the local QHGA and Victorian MDU PHL datasets, along with the global PLSDB database [35] and the ‘AllTheBacteria’ (ATB; [28]) archive using LexicMap (v0.8.0; >60% coverage; [36]), with hits re-annotated using Bakta to ensure consistent feature naming. Additionally, protein homology searches were performed against the NCBI non-redundant database using cblaster (v1.4.0; [37]). Structural conservation was visualised with Clinker (v0.0.32; [38]), and transposition activity was inferred by detecting 6-bp direct target repeats (DRs) at the element’s boundaries. See Supplementary Section S4 for full details.

### Phylogenetic reconstruction and transmission analysis

Core genome clusters were defined using PopPUNK (v2.7.7; [39]) with the *E. faecium* v2 database, and population structure was analysed via the PopPIPE pipeline [40]. Transmission trees were reconstructed using TransPhylo (v1.4.5; [41]) to infer direct and unsampled transmission events from time-labelled phylogenies based on collection dates. Within the primary cluster (44; see Results section ‘Global phylogenomic context and intercontinental transmission’), pairwise alignment-free SNP distances were calculated using SKA (v0.5.0; [42]) and visualised with Seaborn (v0.13.2; [43]). See Supplementary Section S5 for full details.

### Linear plasmid comparative analysis and global screening

To investigate structural evolution, linear plasmid sequences were extracted from 24 LRE from the present study and four surveillance isolates from the state of Victoria, Australia. We performed global synteny analysis using a custom MUMmer (v4.0.1; [44]) workflow: pairwise alignments were generated (`nucmer --maxmatch`) and filtered to retain only 1-to-1 mappings (`delta-filter -1`). A “Global Identity” metric, defined as the total number of matching bases divided by the length of the longest sequence, was used to cluster plasmids via UPGMA (>99.5% identity), with results visualised using pyGenomeViz (v1.6.1; [45]). This clustering was validated by assessing rearrangement distances using Pling (v2.0.1; [46]) and ANI using skani.

To determine the global distribution of this linear plasmid, we screened pELF_LRE_29—representing the most common plasmid structural variant in our outbreak cohort—against our global dataset and the plasmid database PLSDB using LexicMap. High-confidence homologues were identified by applying a minimum query coverage threshold of 70% and alignment length of 2,000 bp. See Supplementary Section S6 for full details.

## Results

### Clonal structure and linezolid resistance in Queensland *Enterococcus*

We characterised 29 linezolid-resistant Enterococcus (LRE) isolates, from 29 patients, collected from public hospitals in Queensland, Australia between June 2023 and June 2024. The cohort was predominantly composed of *E. faecium* (LREfm; *n*=28), with a single detection of *E. faecalis*. Most isolates (25/29) were identified through routine rectal screening, while four were recovered from clinical specimens (Table 1).

**Table 1:**
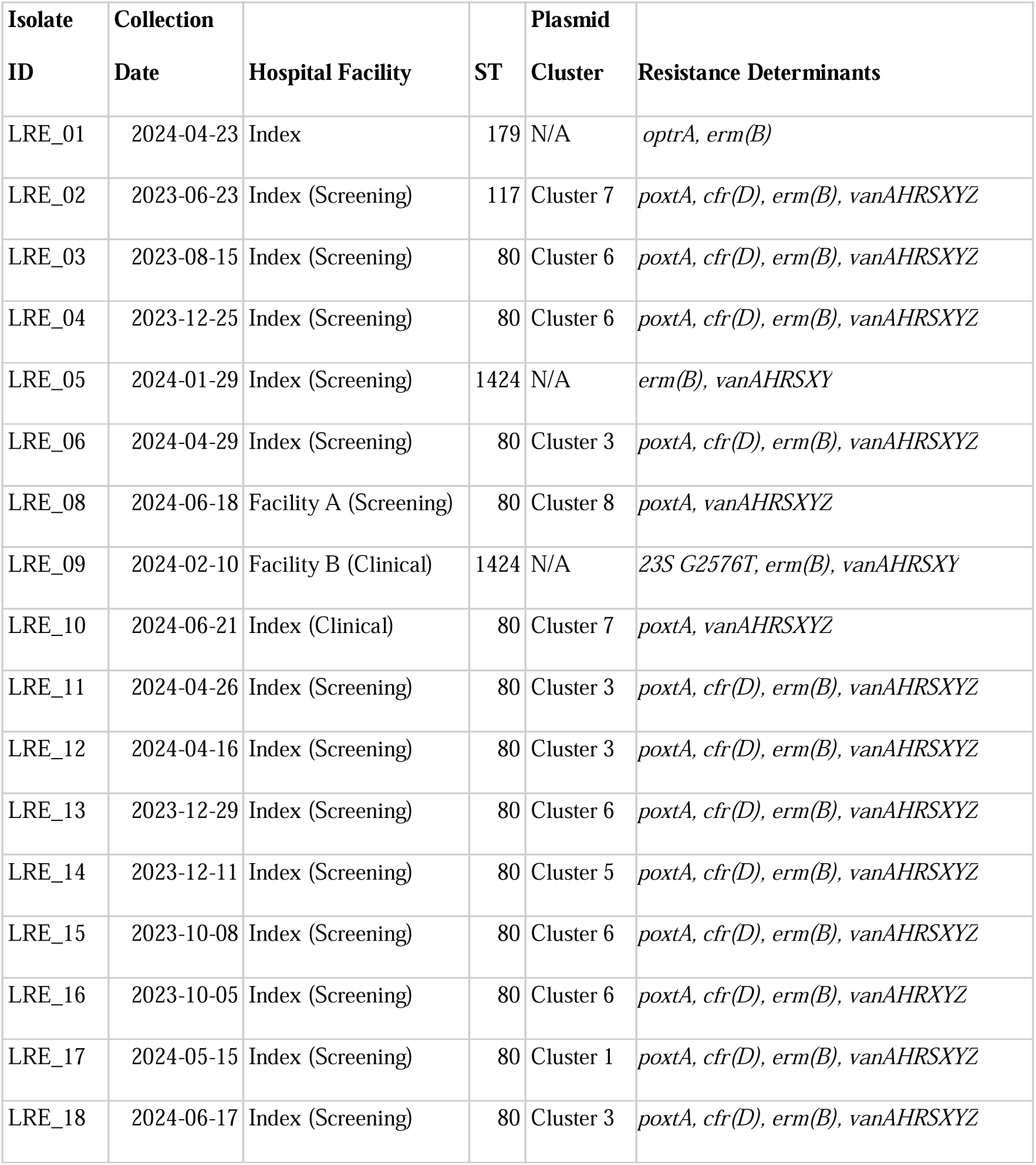
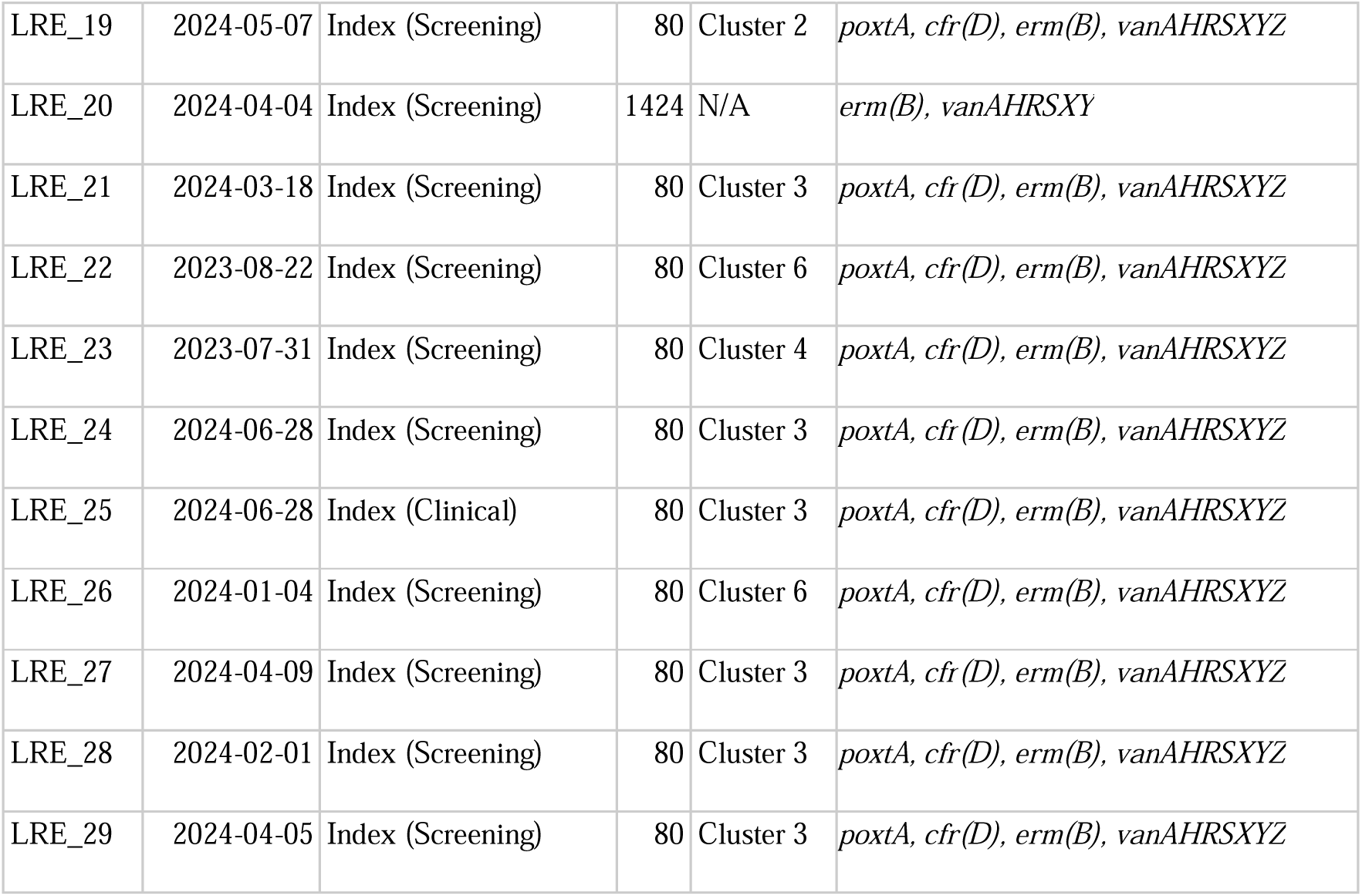
Genomic and epidemiological summary. ”Index” refers to the primary outbreak hospital; “Facility A” and “Facility B” denote anonymized secondary institutions within its referral catchment.

To achieve high-quality genome resolution, all 29 isolates were sequenced with both short-read (Illumina) and long-read (ONT) platforms. However, sample LRE_07 failed to produce a resolved assembly, presenting a highly fragmented contig profile indicative of a mixed or contaminated culture; this sample was consequently excluded from all downstream analyses. All remaining assemblies had 100% completeness and <2.5% contamination (CheckM2; Table S1). Individual isolates harboured between 1 and 10 plasmids, with 24 possessing a linear plasmid. Plasmid sizes ranged from small 1.9 kb elements to large circular plasmids of approximately 251 kb (Table S1).

Multi-locus sequence typing (MLST) revealed that the LREfm population was driven by a single dominant lineage (Table 1). Twenty-three isolates (85%) belonged to ST80 and formed a tight genomic cluster (≤12 SNPs). While this clone was primarily identified at the index hospital, a phylogenetically related isolate was detected at a second facility, within the referral catchment of the index hospital, with associated epidemiology suggestive of inter-hospital dissemination. A smaller cluster of three ST1424 isolates was also detected across two facilities, including a clinical isolate. The remaining LREfm isolate was ST117. See Supplementary Table S2 for full details.

Genomic profiling revealed a multidrug-resistant phenotype across the cohort, predominantly composed of *vanA*- and *poxtA*-positive *E. faecium* (Table 1). While the exact sequence matches the *E. faecium* reference allele *poxtA-Ef* in AMRFinderPlus (protein accession WP_094899500.1), it is functionally equivalent to the staphylococcal *poxtA* prototype [47], despite considerable sequence divergence (72.8% amino-acid identity). For brevity, we use the family symbol *poxtA* throughout the text, though the database-specific *poxtA-Ef* label is retained in some figures and tables to maintain bioinformatic transparency. The single *E. faecalis* (LRE_01), displayed a profile lacking the *vanA* cluster but carrying *optrA*, which confers resistance to linezolid (see Section S7 and Figure S3 for a full description of the *vanA* gene cluster configuration and Table S3 for full resistance profiles).

Linezolid resistance in the *E. faecium* cohort was primarily driven by the clonal expansion of an ST80 lineage (23/27 isolates), alongside a single ST117 isolate (LRE_02). Crucially, resistance in all 24 of these isolates was mediated by the *poxtA* gene—frequently accompanied by *cfr*(D) (21/27)—situated on a near-identical linear plasmid. This vector will hereafter be referred to as an enterococcal linear form plasmid (pELF)-like plasmid (see Section S6.1). Critically, this plasmid was not confined to the ST80 clone: a closely related element was also present in the phylogenetically distinct ST117 isolate (LRE_02), despite the two lineages differing by >2,800 SNPs (see ‘Global phylogenomic context and intercontinental transmission’). The presence of closely related plasmids on highly divergent chromosomal backgrounds points to dissemination of *poxtA*-mediated resistance through plasmid sharing across sequence types, in addition to clonal expansion of ST80—a dynamic we examine in detail below. This plasmid also co-localised the *poxtA* (linezolid) and *vanA* (vancomycin) resistance determinants on a single element. While we did not perform functional transfer experiments, conjugative transfer of pELF-like plasmids has been demonstrated experimentally for related elements [11,48,49]. The genetic architecture observed here therefore raises the possibility that, were this plasmid to mobilise, it could disseminate linezolid and vancomycin resistance concurrently.

In contrast, the ST1424 lineage presented a discordant, plasmid-independent resistance profile. One ST1424 isolate (LRE_09) harboured the 23S rRNA G2576T mutation in three of its six alleles, providing a clear chromosomal basis for resistance, whereas the remaining two (LRE_05 and LRE_20) lacked any known determinants despite exhibiting phenotypic resistance on Vitek 2 (Table S4, Supplementary Section S8).

### Identification of a novel *poxtA*-carrying transposon

The linezolid resistance gene, *poxtA*, was exclusively associated with the pELF-like linear plasmid in all *poxtA*-positive isolates. In addition, sample LRE_02 (ST117) had a second copy of *poxtA* identified on the chromosome.

Examination of the genetic context surrounding *poxtA* revealed a 8185 bp structure distinct from previously described *poxtA*-carrying transposons Tn*6657 and* Tn*6349* [50]. Unlike these transposons, which are bounded by IS*1216E* elements and carry resistance genes *cfr(D)*, *fexB*, and *erm(B)*, the structure identified here is bounded by two IS*1380*-family transposases in the same (direct) orientation. The genetic arrangement comprises an initial IS*1380*-family transposase, followed by a *MerR*-family transcriptional regulator, a GNAT-family N-acetyltransferase, a central transposase, an IS*3*-family transposase, a hypothetical protein, the ABC-F type ribosomal protection protein *poxtA*, and a terminating IS*1380*-family IS*1678* transposase (Figure 1). The upstream IS*1380*-family transposase shared only 67% nucleotide identity with its closest known relative, IS*1678.* As such, we have characterised it as a novel insertion sequence and assigned it the identifier IS*Efa26* (accession PX965924). We also identified 26 bp imperfect inverted repeats (IRs) sharing 22/26 bp identity (IRL: CCTGAATAATTCATAATTTTTCAAAA; IRR: CCTGAATAATTCATACCTTTTATAAA). The downstream IS*1678* element contained 24 bp inverted repeats consistent with reference sequences from ISfinder.

**Figure 1:**
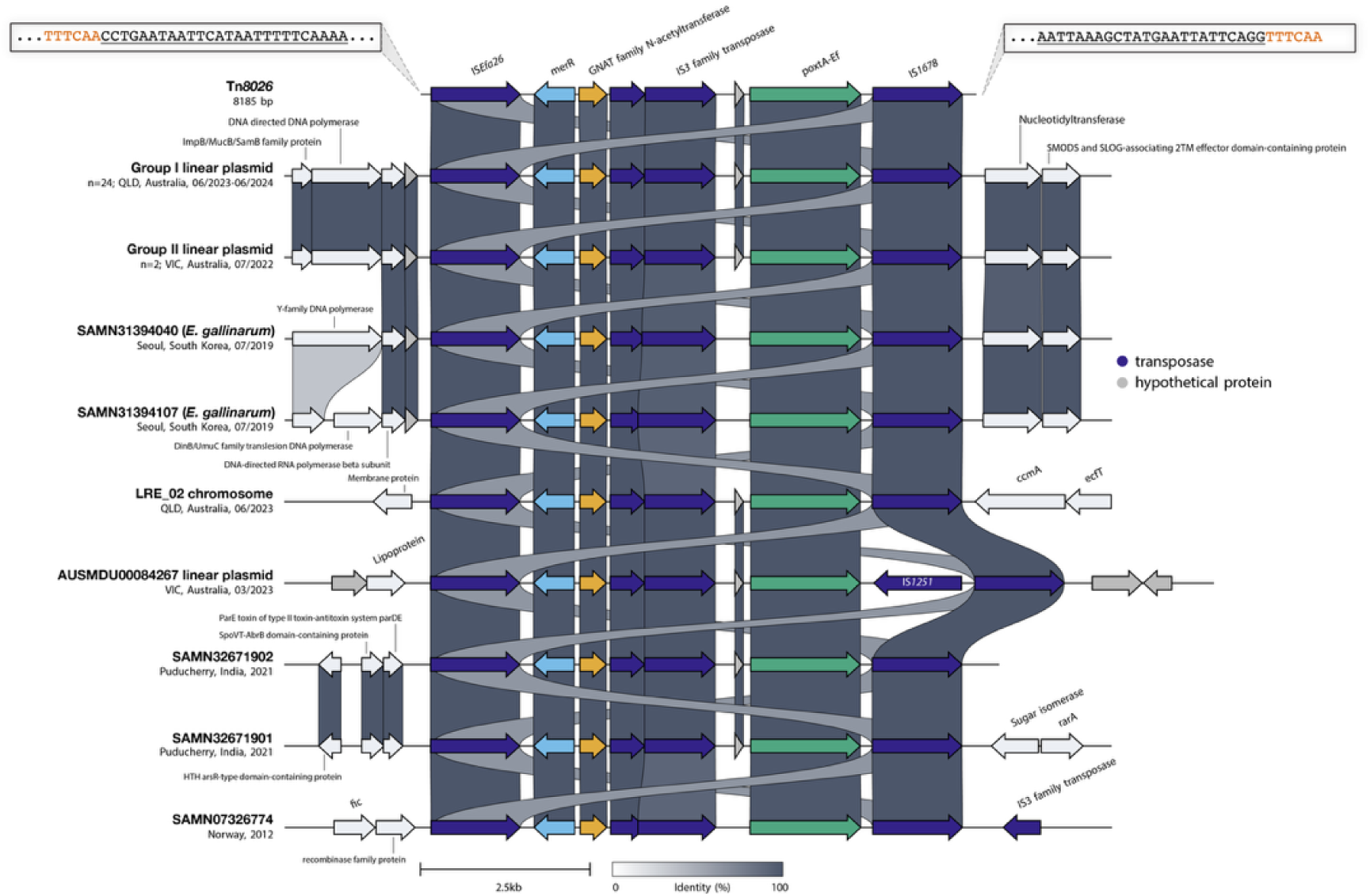
Structural comparison of the novel *poxtA*-carrying transposon Tn*8026*. Alignment of the 8,185 bp composite transposon Tn8026, compared against the linear plasmid-borne copies (Group I; all 24 plasmid-borne copies in this cohort) and the chromosomal copy in LRE_02 (also Group I), recent Victorian isolates (Group II, comprising AUSMDU00074711 and AUSMDU00067942, and the more distantly related AUSMDU00084267), two international isolates identified from the AllTheBacteria archive (SAMN32671901, SAMN32671902), and three isolates identified via BLAST search (the E. gallinarum isolates SAMN31394040 and SAMN31394107, and the Norwegian isolate SAMN07326774). The transposon is bounded by the novel upstream insertion sequence ISEfa26 (left) and the downstream element IS1678 (right). The terminal inverted repeats are represented by the underlined text in the zoom boxes, with the red text showing the direct target DNA sequences. Arrows indicate open reading frames and are oriented in the direction of transcription. Links connecting the tracks represent nucleotide identity between aligned regions, shaded on a colour scale in which darker links denote higher identity. The figure was generated using Clinker [38].

Accordingly, the precise boundaries of this novel composite transposon, designated Tn*8026* (accession PX965923), were defined as the region spanning from the IS*Efa26* IRL to the IS*1678* IRR. Analysis of the regions immediately flanking Tn*8026* revealed the presence of 6 bp direct target DNA sequences (DRs). In the reference isolate LRE_04, the transposon on the linear plasmid was flanked by the DR `TTTCAA`. Crucially, in isolate LRE_02, the plasmid-borne copy of Tn*8026* was flanked by `TTTCAA`, while the chromosomal copy was flanked by a distinct DR (`TCTATT`), providing strong evidence of replicative transposition. A full characterisation of Tn*8026* can be found in Supplementary Section S4

### Global distribution and conservation of Tn*8026*

To determine if Tn*8026* represents a locally emerged element or a globally circulating transposon, we screened the ‘AllTheBacteria’ (ATB) archive [28], containing >2 million bacterial genomes, and genomic datasets from recent Australian hospital surveillance studies in Victoria and Queensland [7,27]. We further interrogated the global diversity of the element using a protein homology search against the NCBI non-redundant protein database with cblaster [37]. Overall, we identified the transposon in 17 external enterococcal isolates, revealing a distribution spanning three continents and multiple species.

Locally, Tn*8026* was detected in three recent clinical LREfm isolates from Victoria, Australia (2022–2023; [7]), confirming its established presence in Australia. Screening of the ATB archive yielded high-confidence hits (>60% coverage) in 11 samples (Table S5). Notably, the complete, intact transposon sequence was recovered from two isolates originating from Puducherry, India (2021; SAMN32671901, SAMN32671902). The remaining nine ATB hits represented fragmented short-read assemblies but confirmed the presence of Tn*8026* in New Zealand (2014), Denmark (2015), Norway (2015–2018), and additional Indian cohorts (2015–2021). Importantly, our cblaster search identified the complete transposon (100% coverage) in a historical isolate from Norway (SAMN07326774) collected in 2012, confirming the element has been circulating in the global population for well over a decade.

We also observed an expansion of the host range beyond *E. faecium*. Complete copies of Tn*8026* were identified in two *Enterococcus gallinarum* isolates from South Korea (2019; [8]), highlighting the transposon’s potential for interspecies transfer.

Comparative genomic analysis revealed convincing structural conservation. An all-vs-all BLASTn analysis demonstrated 100% nucleotide identity between the Tn*8026* reference sequence defined in this study (LRE_04) and the genomes from India, Norway, South Korea, and Victoria (Figure 1). The sole structural deviation was observed in the Victorian isolate AUSMDU00084267, where the transposon backbone was interrupted by an IS*1251* insertion.

### Global phylogenomic context and intercontinental transmission

Next, to understand the broader epidemiology of both our LREfm strains and the pELF-like linear plasmid, we curated a global dataset of 1,880 *E. faecium* genomes comprising the study isolates (LRE; *n*=27), the QGHA (*n*=398; [27]) and Victorian MDU PHL (*n*=379; [7]) cohorts, and globally sourced genomes filtered for high genomic similarity (≥99.8% ANI) or specific resistance profiles (AllTheBacteria *n*=394; GenBank *n*=682). This included all publicly available genomes that were found to carry *poxtA* (*n*=35). Phylogenetic analysis reveals distinct distribution patterns for the linezolid resistance determinants (Figure 2; Table S6). While *cfr* variants are dispersed across diverse clades, *poxtA* carriage is tightly constrained to specific clades, namely, Cluster 44, containing our major LRE ST80 outbreak lineage (*n*=23) and their closest international relatives (*n*=7), and a smaller, distinct ST117 clade. Cluster 44 was found to be the major lineage driving *poxtA*-mediated linezolid resistance and is characterised by ST80 *E. faecium* carrying *poxtA* in a Tn*8026* transposon on a pELF-like linear plasmid.

**Figure 2.**
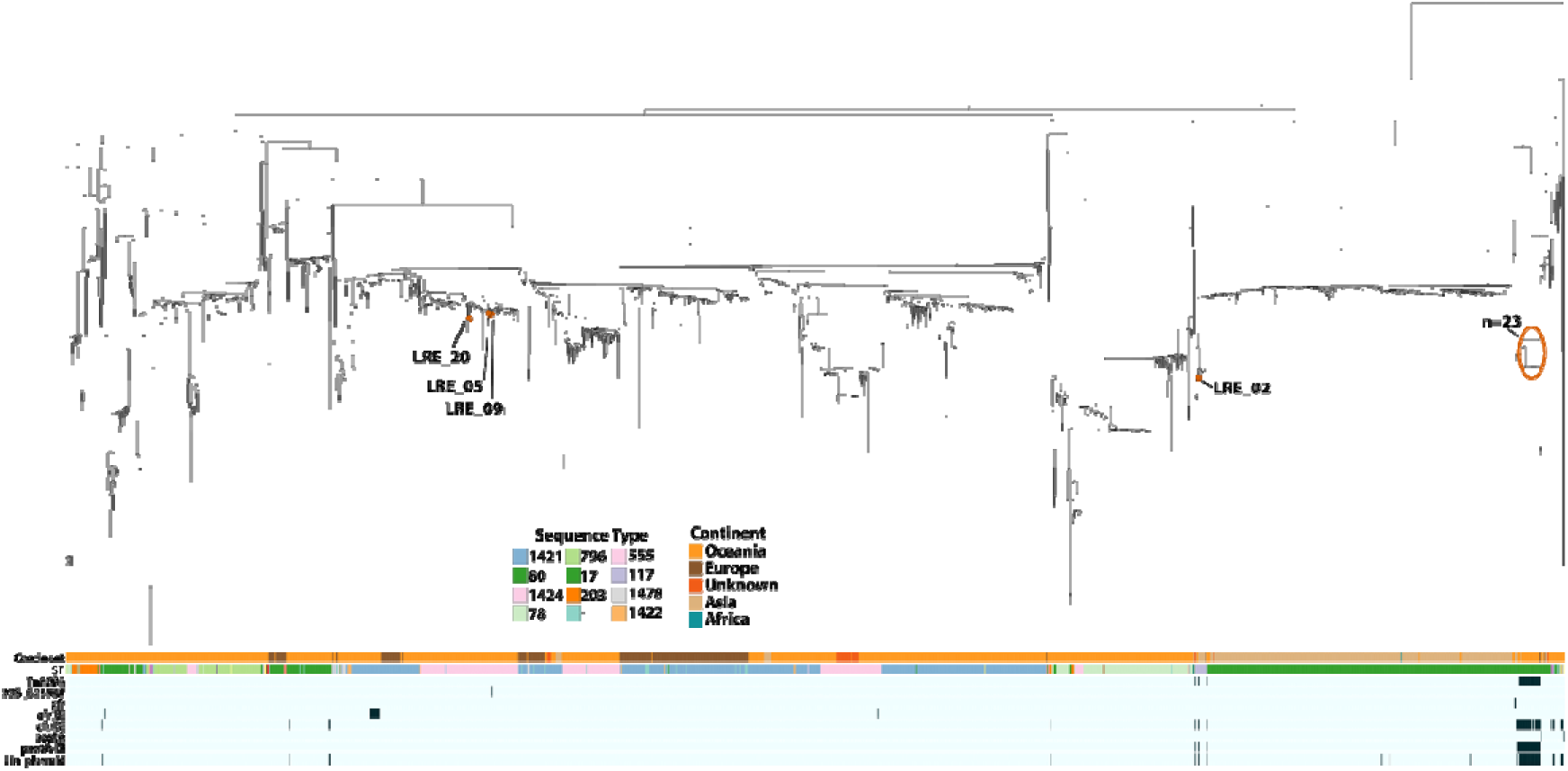
Global population structure of *E. faecium* and distribution of Tn*8026*. A maximum-likelihood phylogeny of 1,880 *E. faecium* genomes was reconstructed using PopPIPE to provide global context for the study isolates (LRE) alongside surveillance cohorts from Queensland (QGHA) and Victoria (MDU), and globally sourced genomes from AllTheBacteria and GenBank. The metadata heatmaps (bottom) display the continent of origin, sequence type, Tn*8026* and linear plasmid presence (black indicates presence), and linezolid resistance determinants (presence of *poxtA*, *poxtA-Ef*, *cfr*, *cfr(B), cfr(D),* and 23S mutation G2576T). The study isolates are annotated with red points, with those not explicitly named being present in the n=23 group circled. An interactive version of this tree can be accessed at https://microreact.org/project/keKHSzjhW725GManpizMTM-efm-poppipe.

Separately, the ST117 group includes study isolate LRE_02, a Victorian isolate (AUSMDU00084267, 303 SNPs), and an Indian isolate (SAMN32671898, 47 SNPs)—all of which harbour the linear plasmid and Tn*8026* element. To contextualise the imported ST117 lineage, we examined its phylogenetic neighbourhood. The closest genomic matches to LRE_02 were recent clinical isolates from India (April–May 2024; ∼54 SNPs) and Singapore (December 2023–May 2024; 57–75 SNPs); however, notably, none of these closely related isolates harboured the linear plasmid, except SAMN32671898. The Victorian surveillance case remained distantly related (303 SNPs). Broader screening identified only two other isolates near LRE_02 in the global phylogeny that possessed the linear plasmid: the aforementioned Indian isolate (SAMN32671898, 2021) and a Norwegian isolate (SAMEA115490957, 2018).

Isolate LRE_02 (ST117) was recovered in June 2023 from a patient recently hospitalised in India for major surgery. During this admission, the patient shared a ward with the individual who subsequently yielded the index ST80 outbreak isolate, LRE_23, in July 2023. While separated by 2,821 SNPs, both isolates harbour the highly conserved linear plasmid carrying Tn*8026*. Furthermore, functional evidence of the transposon’s mobility was observed in the imported ST117 isolate (LRE_02), where Tn*8026* was identified integrated into the chromosome in addition to its presence on the linear plasmid vector.

Turning to the clonal ST80 outbreak, Bayesian transmission reconstruction, shown in Figure 3, estimated the Cluster 44 index case to circa 2014 (95% credible interval: Oct. 2013 – Jan. 2014), postdating the global emergence of Tn*8026* (Norway, 2012). The model inferred a high-probability introduction of the lineage from the Indian subcontinent to Victoria, supported by genomic proximity (11 SNPs; Section S9) and documented patient travel history [7]. This imported lineage was subsequently linked to the Queensland LRE cohort via an indirect transmission path involving two unsampled intermediate hosts (29 SNPs), consistent with undetected interstate dissemination prior to outbreak detection. However, the epidemiological link between the imported ST117 (LRE_02) and the ST80 index case (LRE_23) adds critical context to this model, with potential transmission scenarios explored further in the Discussion.

**Figure 3.**
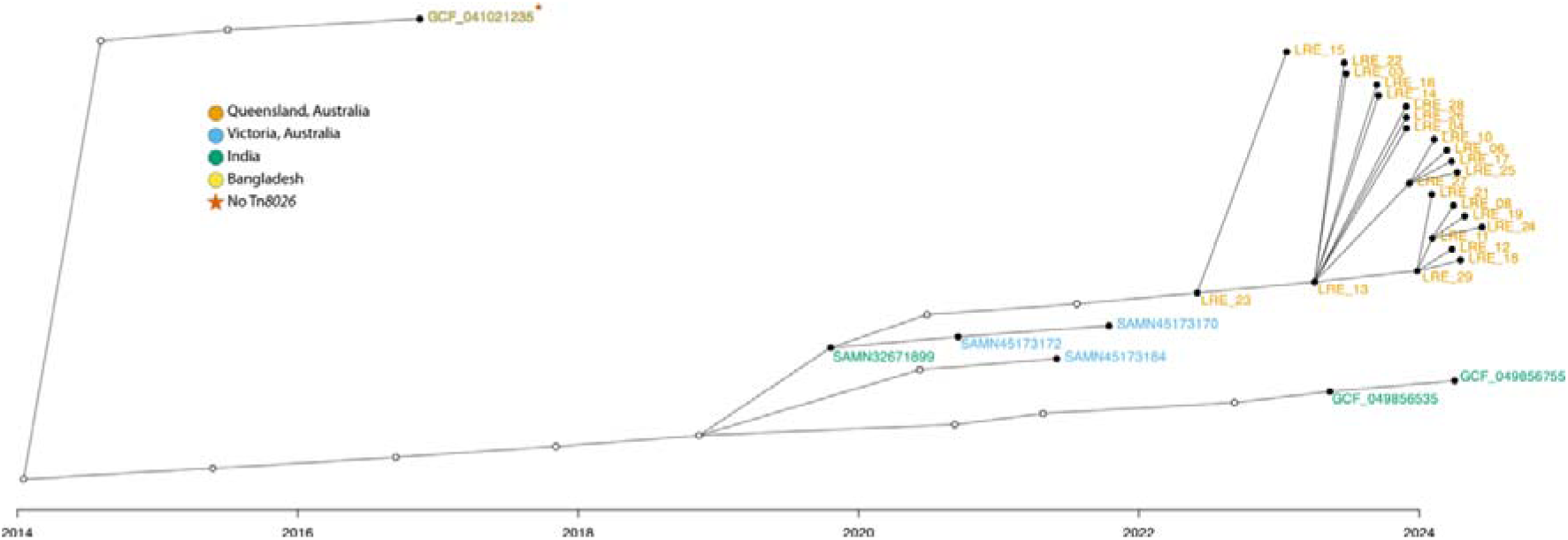
Consensus transmission tree of the Tn*8026*-positive *E. faecium* ST80 Cluster 44. This visualisation represents the transmission history reconstructed using TransPhylo, where the x-axis indicates the estimated date of infection for each host and the y-axis is arbitrary. Individual hosts are represented by nodes, where filled circles indicate sampled isolates, coloured by geographic origin, and unfilled circles represent inferred unsampled intermediate hosts (missing links) required by the model to explain the transmission chain. Edges indicate inferred transmission events.

### Linear plasmid architecture and global dissemination

A defining feature of these assemblies was the presence of plasmids with linear topology in 24 of the 28 isolates. These pELF-like plasmids [12] failed to circularise during assembly and ranged in size from 108 kb to 133 kb (Table S2; [1]). Each linear plasmid has a hybrid topology: a hairpin end, and an “open” end, which does not fold back on itself and has a terminal protein covalently attached to the 5’ end (Figure S6). This matches the findings of Beh *et al.*, who recently found 4 LREfm pELF-like plasmids with the same topology in Victoria [7].

Global synteny analysis partitioned the 28 linear plasmids (24 from this study and 4 from Victoria) into 10 clusters, revealing a dominant ∼132 kb lineage and distinct deletion variants (Figure 4; Section S10). Most plasmids (*n*=23) belonged to a highly conserved “Core LRE Complex” (Clusters 1–6; Table 1) characterised by a full complement of resistance and cargo genes. Structural diversification within the cohort was driven by presumed sequential deletion events at the “left” (hairpin) end of the plasmid.

**Figure 4.**
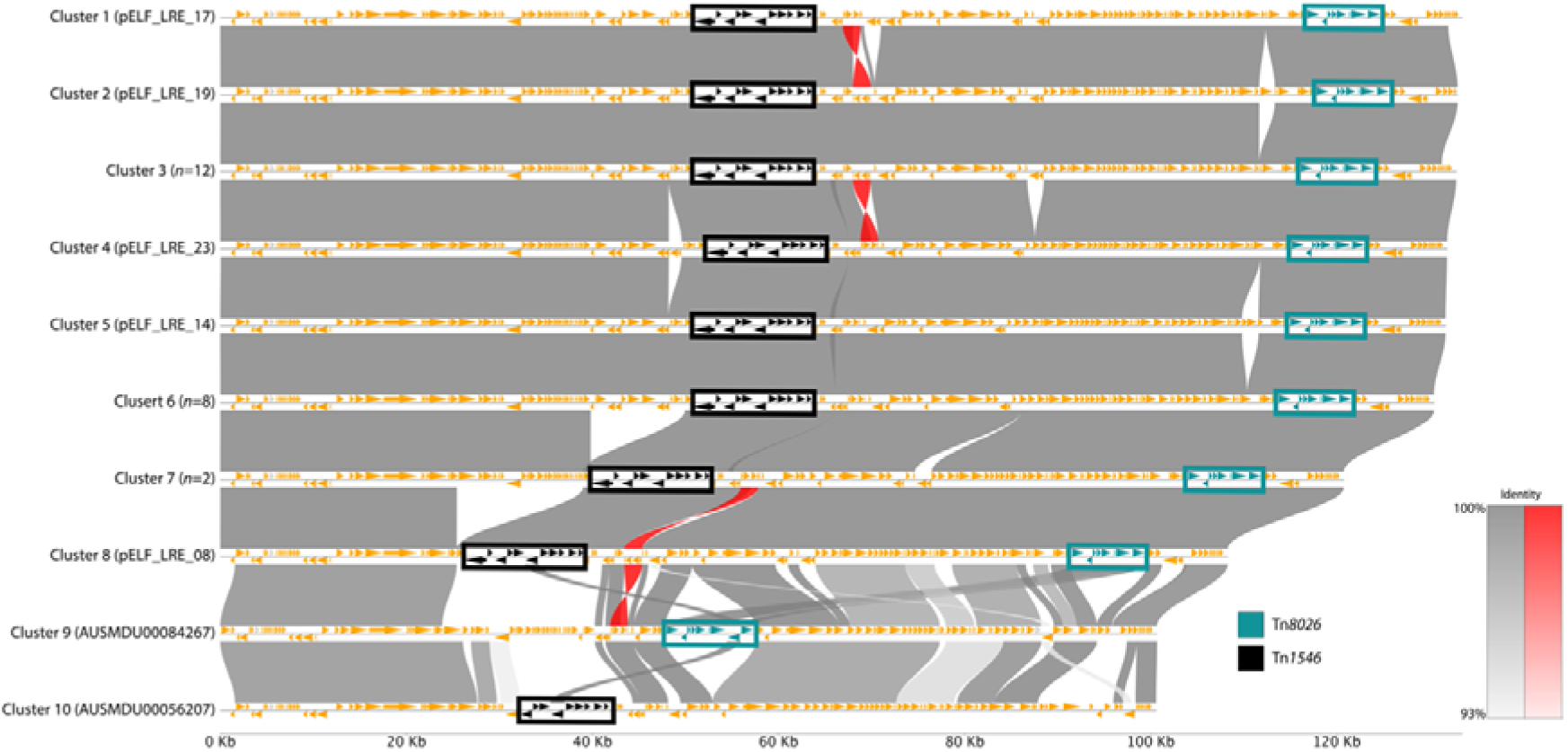
Synteny analysis of representative linear plasmids. Pairwise nucleotide alignments between the ten representative linear plasmids were generated using MUMmer and visualised with pyGenomeViz. Plasmids are ordered by cluster number (1–10), corresponding to decreasing length. The grey and red bands connecting genomes represent regions of conserved synteny, with colour intensity indicating percent identity, and red indicating inversion. Orange arrows represent annotated genes. Key resistance elements are highlighted with coloured boxes: Tn*1546* (vancomycin resistance) in black and Tn*8026* (linezolid resistance) in teal. The core LRE lineage (Clusters 1–6) shows high structural conservation. Large deletions are evident in Clusters 7 and 8, visible as gaps in the alignment bands relative to the longer plasmids. Clusters 9 and 10 display significant structural divergence, particularly in the presence and location of Tn*1546* and Tn*8026*.

The most significant variation was observed in Cluster 7 (pELF_LRE_10 and pELF_LRE_02), where a ∼10.6 kb region containing the *cfr*(D) and *erm*(B) resistance genes was lost. A further reductive event was identified in the singleton Cluster 8 (pELF_LRE_08), which represents a smaller variant (∼108 kb) lacking both the *cfr(D)*/*erm(B)* module and an additional ∼13.3 kb of cargo. These stepwise deletions, spanning ∼10.6–23.9 kb relative to the core lineage and consistently localised to the region upstream of Tn*1546* (Figure 4), are detailed in Section S10. The remaining two Victorian isolates formed distant singleton clusters with divergent resistance region architectures, confirming that while the linear backbone is circulating regionally, the specific drug resistance cargo varies.

Screening of the global dataset with LexicMap identified the pELF-like linear plasmid backbone in 45 unique genomes across Australia, China, India, and Norway (Section S10), whereas querying PLSDB returned no hits. While the backbone itself was detected in historical Queensland isolates (2019), structural analysis demonstrated only 72-78% coverage. Conversely, the Indian isolate SAMN32671899 (2021) carried a pELF-like variant that shared higher breadth of coverage with our outbreak pELF-like linear plasmid (81.9%) and contains Tn*8026,* strongly supporting an independent importation event for the 2023 outbreak lineage. Overall, this aligns with the transmission reconstruction, showing a stepwise increase in plasmid identity along the importation route from India to Victoria (80.7% coverage) and finally to the 2023 LRE cohort (Cluster 6; 93.8%), consistent with the importation of a specific LRE lineage, rather than the re-emergence of a local historical variant.

Strikingly, alignment of the two Tn*8026*-positive *E. gallinarum* isolates from South Korea against the outbreak reference plasmid (pELF_LRE_29) revealed ∼90.9% sequence coverage (Section S6). This confirms that the pELF-like linear plasmid, complete with its linezolid resistance cargo, has successfully mobilised across enterococcal species boundaries.

## Discussion

This study characterises the genomic epidemiology of a linezolid-resistant *E. faecium* (LREfm) outbreak driven by the importation of an ST80 clone carrying a novel transposon, Tn*8026*, on a linear plasmid. While linezolid resistance in *E. faecium* has historically been dominated by *de novo* chromosomal mutations under antibiotic pressure [51,52], our findings highlight the role of transferable resistance mechanisms in the global dissemination of AMR. We provide strong genomic evidence that the resistance determinant *poxtA* was not acquired locally but was imported into Australia from the Indian subcontinent via an enterococcal linear form plasmid (pELF) vector that is difficult to resolve using standard genomic surveillance.

Integrating clinical epidemiology with high-resolution genomics reveals a complex transmission dynamic preceding the ST80 outbreak. The recovery of the ST117 isolate (LRE_02) from a patient recently hospitalised in India, who subsequently shared a ward with the ST80 index case (LRE_23), establishes a clear importation event linking two distinct lineages. Intriguingly, both isolates harbour the linear plasmid. When examining the whole genome, the closest relatives to LRE_02 by absolute genetic distance are recent isolates from India (54 SNPs) and Singapore (57–75 SNPs); however, these highly related strains lack the linear plasmid. Conversely, the closest phylogenetic neighbour to LRE_02 is a Victorian surveillance isolate (AUSMDU00084267) separated by 303 SNPs. While this Victorian isolate does carry the linear plasmid, its plasmid exhibits a substantially different structure (Cluster 9). Broader screening identified only two other isolates near LRE_02 in the global phylogeny that possess the linear plasmid (India 2021 and Norway 2018). Because the most genetically similar isolates lack the plasmid, and the nearest plasmid-bearing relative is comparatively distant and structurally divergent, the exact sequence of plasmid acquisition in this cohort remains ambiguous. This convergence could be explained by an *in vivo* horizontal transfer from the imported ST117 strain into a locally circulating ST80 lineage within the ward, the co-importation of both plasmid-bearing strains from India, or independent plasmid acquisition by both lineages from an unsampled local reservoir. Although the precise sequence of events cannot be definitively resolved without exhaustive sampling of intermediate hosts and environmental reservoirs, this epidemiological link and our Bayesian reconstruction strongly underscore the capacity of these mobile elements to move undetected across borders and between distinct bacterial populations.

A defining feature of this cohort was the localisation of the MDR determinants on a linear plasmid. These pELF-like replicons have recently emerged as critical vectors for *vanA*, *optrA*, and *poxtA* in *E. faecium* populations [7,11,48]. Crucially, the identification of these plasmids is frequently hampered by the limitations of short-read sequencing, which often fragments linear replicons due to their complex terminal structures and repetitive elements [53]. By employing a hybrid assembly approach, we resolved the complete structure of the outbreak plasmid, revealing it to be a highly conserved vehicle for Tn*8026*. Interestingly, while we detected the linear plasmid backbone in historical Queensland isolates from 2019 [27], the specific outbreak plasmid carried a distinct variant of Tn*8026* that matched an Indian isolate (SAMN32671899) more closely than the local historical variants. This suggests that the linear backbone serves as a receptive, globally distributed platform capable of acquiring diverse MDR modules, effectively ‘upgrading’ local lineages or facilitating the movement of imported resistance mechanisms.

The reconstruction of the transmission chain offers high-resolution insight into the movement of this lineage. Bayesian transmission analysis inferred a high-probability linkage descending from the Indian subcontinent lineage to the Queensland LRE cohort. The analysis indicated the presence of unsampled intermediate hosts separating the Indian isolate from the Queensland outbreak strain (LRE_23) by a genetic distance of 29 SNPs. The location of these unsampled intermediates—whether in India, Australia, or a third region—remains unknown, highlighting the challenges of tracking mobile lineages through gaps in global surveillance. However, the close genomic proximity (11 SNPs) of related Victorian isolates with documented travel history to the region provides strong support for an importation hypothesis. This silent dissemination is concerning, as it implies the Tn*8026* lineage was circulating undetected prior to the clinical outbreak, likely facilitating the establishment of this resistance mechanism in the local hospital network.

The discovery of Tn*8026* provides insight into the modular evolution of *poxtA*-mediated resistance. Structurally, the element is defined by the novel insertion sequence IS*Efa26* and an IS*1678* element, rather than the IS*1216E*-driven modules typically associated with *poxtA* dissemination [50]. Our global screening identified this transposon in a historical Norwegian isolate from 2012, indicating that the mechanism was established in the global gene pool well before the current outbreak. Several genomic signatures indicate that Tn*8026* has likely been mobile: it is bounded by 6-bp direct target-site duplications (Section S4), a characteristic product of transposition that is not generated by homologous recombination [54], and the same element occurs in multiple independent contexts—chromosomal in LRE_02 as well as plasmid-borne, and at a different plasmid position in a Victorian isolate (AUSMDU00084267). While we did not assay for active transposition (e.g. circular intermediates), these signatures are consistent with an element that has been mobile over its evolutionary history. Furthermore, our discovery that the two hospital-associated *E. gallinarum* isolates [8] harbour not only Tn*8026* but the pELF-like linear plasmid (>90% coverage) demonstrates that this specific multidrug-resistant vector is capable of crossing species boundaries. While currently rare, this capacity for interspecies mobility underscores the need for continued surveillance of Tn*8026* across the enterococcal genus.

Our synteny analysis revealed a likely pattern of stepwise reductive evolution within the linear plasmid cohort. We observed sequential deletions of large MDR regions (containing *cfr*(D) and *erm(B)*) in specific sub-clusters (Clusters 7 and 8). These plasmid-level structural rearrangements appear to be mediated by IS*1216E*, which flanked the deleted regions. We hypothesise that this ‘genome trimming’ represents a fitness adaptation. Large linear plasmids can impose a metabolic burden on the host [15]; by shedding extra cargo genes while retaining the essential linear backbone and core resistance determinants, the plasmid may stabilise its relationship with the host in the absence of specific antibiotic selection pressures. This phenomenon mirrors findings by Sun *et al.*, who described similar IS-mediated reductions in *vanM* plasmids to restore host fitness [55].

The reliance on routine phenotypic testing for surveillance means that while the presence of resistance is identified, the route of transmission often remains obscured. While standard short-read whole-genome sequencing is typically highly effective for resolving these transmission networks, this study demonstrates that it is insufficient for monitoring the spread of linear plasmids. Future surveillance strategies must therefore incorporate long-read sequencing or specific bioinformatic workflows to detect and track these atypical linear replicons. A limitation of our study is the lack of community sampling; the unsampled intermediates inferred by our models likely resided in reservoirs that remain largely opaque to hospital-based surveillance.

In conclusion, we describe the importation and dissemination of a linezolid-resistant *E. faecium* lineage carrying the novel transposon Tn*8026* on a linear plasmid. Crucially, we demonstrate that this vector is actively crossing enterococcal species boundaries. These findings underscore the role of atypical linear plasmids as vehicles for multidrug resistance, highlighting the necessity of advanced long-read genomic surveillance to detect and characterise these complex mobile genetic elements.

## Data Availability

Raw sequencing reads (Illumina and ONT) and hybrid assemblies generated in this study have been deposited in the European Nucleotide Archive (ENA) under BioProject PRJEB106135. Individual sample accession numbers are listed in Table S8.
The novel mobile genetic elements described in this study have been assigned designations Tn*8026* and IS*Efa26* by us. These identifiers were selected to adhere to standard nomenclature conventions while avoiding conflict with recent allocations. Formal requests for accession numbers were submitted to the Transposon Registry and ISfinder; however, as no response was received at the time of publication, these names were assigned to facilitate consistent reporting. We remain committed to formally registering these elements with the respective databases once communication is established. We have submitted sequences to the ENA with accessions PX965923 (Tn*8026*) and PX965924 (IS*Efa26*).
Detailed notes of the manual resolution of linear plasmid assemblies has been archived in Zenodo (www.doi.org/10.5281/zenodo.18626625).

https://doi.org/10.5281/zenodo.18626624

## Data Summary

Raw sequencing reads (Illumina and ONT) and hybrid assemblies generated in this study have been deposited in the European Nucleotide Archive (ENA) under BioProject PRJEB106135. Individual sample accession numbers are listed in Table S8.

The novel mobile genetic elements described in this study have been assigned designations Tn*8026* and IS*Efa26* by us. These identifiers were selected to adhere to standard nomenclature conventions while avoiding conflict with recent allocations. Formal requests for accession numbers were submitted to the Transposon Registry and ISfinder; however, as no response was received at the time of publication, these names were assigned to facilitate consistent reporting. We remain committed to formally registering these elements with the respective databases once communication is established. We have submitted sequences to the ENA with accessions PX965923 (Tn*8026*) and PX965924 (IS*Efa26*).

Detailed notes of the manual resolution of linear plasmid assemblies have been archived in Zenodo [1].

## Impact statement

Linezolid is a critical last-resort antimicrobial for treating multidrug-resistant enterococcal infections, and the emergence of transferable resistance poses a severe threat to clinical management. By investigating a recent hospital-associated outbreak of linezolid-resistant *Enterococcus faecium* in Queensland, Australia, we identified that resistance was driven by a novel transposon (Tn*8026*) carrying the *poxtA* gene. Crucially, long-read sequencing revealed this transposon is situated on a linear plasmid. By screening global genomic datasets, we traced the evolutionary trajectory of this mobile genetic element, demonstrating intercontinental spread from the Indian subcontinent to Australia and revealing a global reservoir that has circulated undetected since at least 2012. This study represents a step advance in our understanding of the enterococcal mobilome by proving that atypical linear plasmids act as systematically overlooked, global drivers of multidrug resistance. These findings are of broad utility to genomic epidemiologists, bioinformaticians, and clinical microbiologists, emphasising the essential role of long-read genomic surveillance in detecting and resolving elusive resistance vectors before they establish endemicity in hospital networks.

## Acknowledgements

This work was supported by resources provided by The University of Queensland Research Computing Centre’s Bunya supercomputer [56], with funding from The University of Queensland, Brisbane, Australia.

## Author Contributions

**Michael B. Hall**: Conceptualisation, Methodology, Software, Formal Analysis, Investigation, Data Curation, Writing – Original Draft, Visualisation, Writing – Review and Editing. **Yiyang Xue**: Formal Analysis, Investigation. **Tricia S. E. Lee**: Investigation, Resources. **Ella Herring**: Investigation, Resources, Writing – Original Draft. **Jocelyn Hume**: Investigation, Resources. **Ryan R. Wick**: Methodology, Software, Validation, Writing – Review & Editing. **Tim Kidd**: Investigation, Resources. **Naomi Runnegar**: Resources, Investigation, Data Curation, Writing – Review & Editing. **Patrick N. A. Harris**: Conceptualisation, Resources, Writing – Review & Editing, Supervision. **Bianca Graves**: Conceptualisation, Investigation, Resources, Data Curation, Writing – Original Draft, Writing – Review & Editing, Supervision. **Leah W. Roberts**: Conceptualisation, Methodology, Resources, Writing – Original Draft, Writing – Review & Editing, Supervision, Funding acquisition.

## Ethics Statement

This work was conducted under the oversight of the Coronial and Public Health Sciences Human Ethics Committee (HREC/17/QFSS/6) and The Queensland Children’s Hospital Human Research Ethics Committee (HREC/22/QCHQ/85249), with a waiver for individual patient consent. All data in the manuscript and supplementary files are fully de-identified.

