## Supplementary Material for "Novel transposon Tn*8026* contributes to the global spread of transmissible linezolid resistance in *Enterococcus* via a linear plasmid"

### Supplement

#### S1. Isolate sequencing

Short read sequencing was performed on Illumina MiniSeq (150 bp paired-end) using the Illumina DNA Prep library prep kit. Surveillance isolates initially triggered interest due to confirmed phenotypic linezolid resistance. Genomic screening and cluster identification were undertaken using our custom, in-house microbial genomic analysis pipeline, SnapperRocks (https://github.com/FordeGenomics/SnapperRocks), which implements Nesoni (https://github.com/Victorian-Bioinformatics-Consortium/nesoni) and CATHAI [1] for genomic clustering. Genotypic resistance profiles were confirmed using a combination of LRE-Finder 1.0 [2] and NCBI AMRFinderPlus (v3.12.8; [3]). Based on these screening criteria, all cohort isolates were subsequently selected for long-read sequencing via Oxford Nanopore Technologies (ONT). This long-read sequencing was deployed specifically to resolve the complete genomic architecture and exact structural context (e.g., chromosomal versus plasmid integration) of the identified linezolid resistance determinants.

Pure single colonies were collected from overnight purity plates grown at 37°C on horse blood agar and inoculated into 5mL of 1x LB Broth (Gibco™) and grown at 37°C overnight with 250 rpm shaking. 1mL of overnight culture was pelleted and nucleic acid extracted using the QIAamp® PowerFecal® Pro DNA Kit (QIAGEN), as per manufacturer's recommendations. DNA was quantified using a Qubit 4 Fluorometer (Q33238) with the dsDNA BR Assay Kit (Q32853). A total of 200ng of each extract was prepared and all 29 samples were pooled using the Rapid Barcoding Kit 96 V14 (SQK-RBK114.96) and sequenced on a PromethION 2 Solo (Oxford Nanopore Technologies) using a R10 flow cell for up to 48 hours. Raw ONT data were basecalled with Dorado (v1.1.1) using the v5.2.0 super-accuracy (SUP) model.

#### S2. Genome assembly

We generated long read–only assemblies using Autocycler (v0.5.1; [2]) after removing reads shorter than 1000 bp. We executed Autocycler using the pipeline outlined at https://github.com/rrwick/Autocycler/tree/main/pipelines/Slurm_Autocycler_Bash_script_by_Michael_Hall. This script uses Autocycler's assembly helper script, which provides a genome size estimation generated by LRGE (default settings; v0.2.1; [5]) and exclude contigs with a depth <10% of the longest contig (`--genome_size <size> --min_depth_rel 0.1`) for all assemblies. The assembler tools used were Flye (v2.9.6), Canu (v2.3; [6]), Raven (v1.8.3; [7]), Myloasm (v0.5.1; [8]), metaMDGB (v1.3.1; [9]), Miniasm (v0.3; [10]), NECAT (v0.0.1; [11]), NextDenovo (v2.5.2; [12]), and Plassembler (v1.8.2; [13]).

Samples containing a linear plasmid required additional manual curation [14]. For these, we visualised the Autocycler assembly graphs in Bandage (v0.9.0; [15]) and resolved divergent paths by removing the lowest-depth node, prioritising retention of the longest continuous sequence at the open (protein-capped) end of the plasmid.

To maximise recovery of linear plasmid termini—which are often better represented in short-read data [16]—we reassembled samples containing linear plasmids with Unicycler (v0.5.1; [17]) using both long and short reads. Unicycler was run in bold mode, with the Autocycler assembly supplied as the existing long-read assembly. When Unicycler produced a longer linear plasmid than Autocycler, we adopted the Unicycler version; otherwise, we retained the original Autocycler plasmid. In 21/24 of the linear plasmids, Unicycler lead to an increase in sequence length (mean 725 bp, range 35–3114 bp; Table S2 and Figure S1). Circular replicons were reoriented to begin with the relevant replication initiation gene with dnaapler (v1.3.0; [18]).

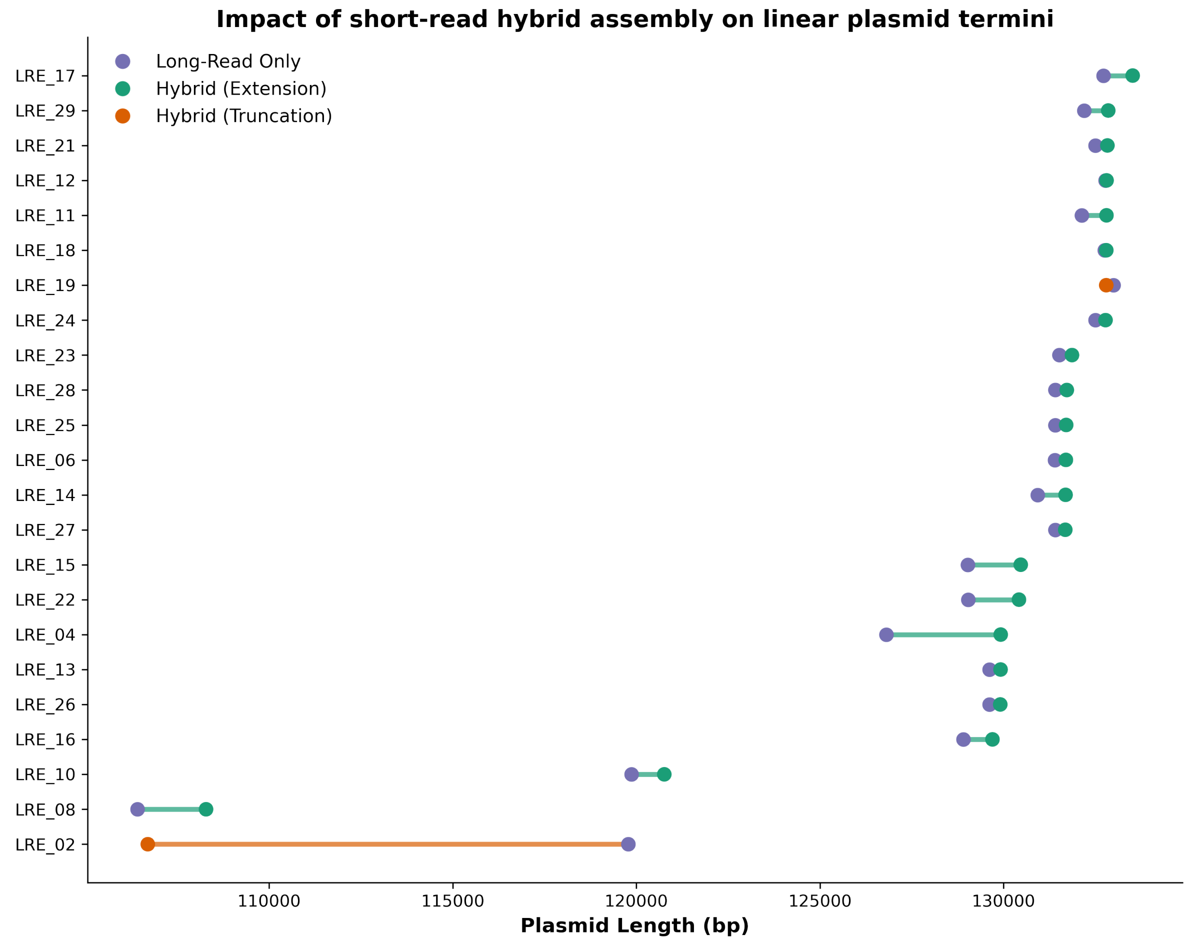


**Figure S1: Impact of short-read hybrid assembly on linear plasmid terminal length.** Blue points indicate the length (x-axis) of each sample's (y-axis) linear plasmid from the long read-only Autocycler assembly. The blue points are connected to points that show the length after reassembling the linear plasmid with short and long reads in Unicycler, using the long read-only assembly as the starting point, with green points and connecting lines indicating an increase in length and orange indicating truncation.

All assemblies were polished sequentially with long, then short, reads. Long-read polishing was performed using Medaka (v2.1.0) with the `--bacteria` option. The resulting assemblies were then polished with Illumina data using Polypolish (v0.6.0; default options; [19]), followed by Pypolca (v0.6.0; [20]) with the `--careful` option.

#### S3. Global genomic dataset curation

To place the *E. faecium* study isolates (LREfm cohort; n=27) into a broader epidemiological context, a background dataset was compiled from multiple public sources. We incorporated genomic data from a Queensland Genomics Health Alliance (QGHA) study (BioProject PRJNA797179; n=398 successfully downloaded of 413 *E. faecium* genomes; [21]) and surveillance isolates from the Microbiological Diagnostic Unit Public Health Laboratory (MDU PHL, Victoria; n=379; [22]). To maximise the detection of related transmission events and resistance reservoirs, we also mined the 'AllTheBacteria' (ATB) archive (n=36,572 *E. faecium*; [23]) and NCBI GenBank/RefSeq (n=40,281 *E. faecium*; [24,25]).

*E. faecium* genomes from ATB and GenBank/RefSeq were screened for daptomycin and linezolid resistance markers using AMRFinderPlus (v4.0.23; database version 2025-07-16.1; [3]). Daptomycin resistance was defined by the presence of known mutations in *liaR* (e.g., W73C), *liaS* (e.g., T120A), or *cls* family genes, while linezolid resistance was defined by the presence of *poxtA* (and its homologues), *optrA*, *cfr* variants, or 23S rRNA mutations (G2576T) with >90% coverage. To identify phylogenetically relevant isolates regardless of resistance status, we calculated Average Nucleotide Identity (ANI) between our LRE isolates and the public datasets using skani (v0.3.1; [26]). We built a sketch database (`skani sketch` [default parameters]) and searched each public genome (`skani search`) against all LRE isolates, retaining only pairs where the aligned fraction exceeded 80% for both genomes (`--both-min-af 80`) and ANI was ≥99.8%. After removing 154 ATB genomes already represented in the QGHA or MDU datasets, 2,918 unique ATB BioSamples were retained; after removing 468 GenBank/RefSeq genomes already represented in ATB, QGHA, or MDU, and applying RefSeq-preference deduplication for assemblies from the same BioSample, 682 unique GenBank assemblies remained. To correct for sampling bias, daptomycin- and linezolid-susceptible isolates were randomly subsampled to match the count of resistant isolates within each source (ATB: 197 resistant, 197 subsampled susceptible; GenBank: 341 resistant, 341 subsampled susceptible). Combined with the 398 QGHA isolates, 379 MDU isolates, and the 27 LRE study isolates, the final dataset comprised 1,880 genomes (see Figure S2).

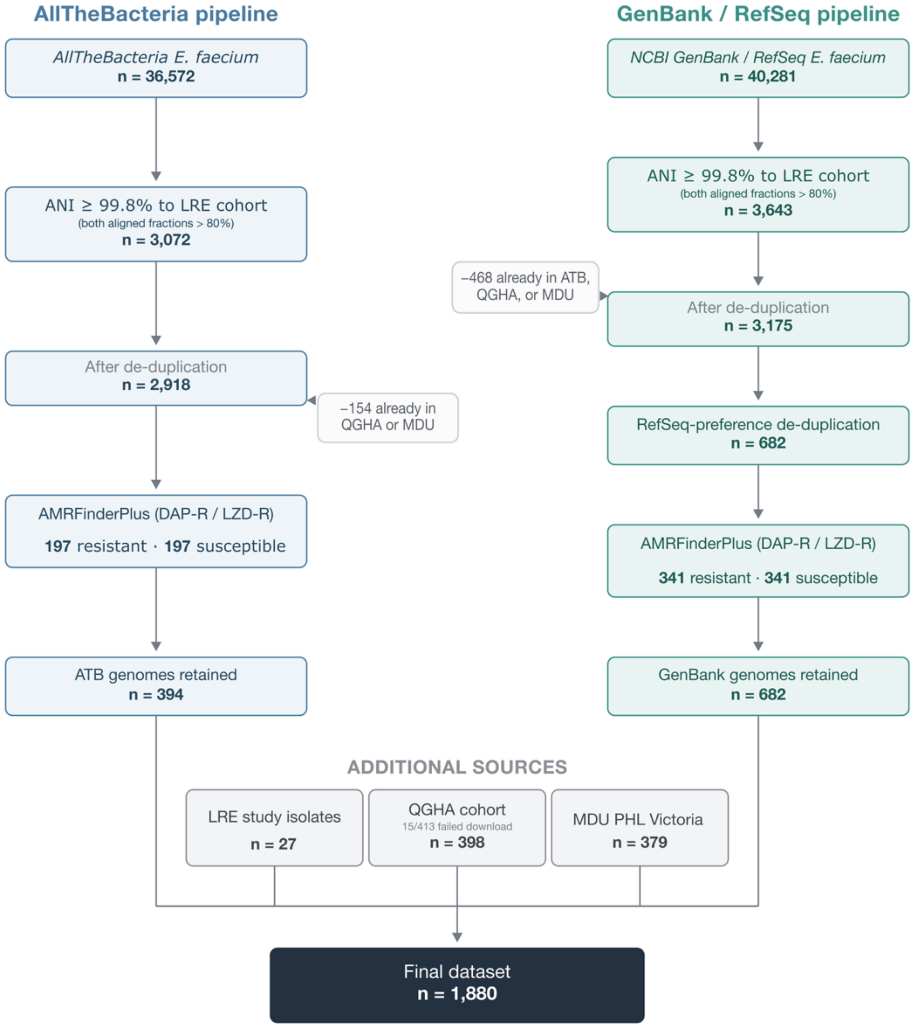


**Figure S2: Construction of the global Enterococcus faecium genomic dataset.** Flowchart illustrating the stepwise curation of the 1,880-genome background dataset used for phylogenetic and epidemiological analyses. Two parallel pipelines sourced genomes from AllTheBacteria (ATB; left) and NCBI GenBank/RefSeq (right), each filtered by Average Nucleotide Identity (ANI ≥99.8%, minimum aligned fraction >80% for both genomes) relative to the LRE study cohort using skani. Overlapping genomes already represented in the QGHA or MDU datasets were removed prior to resistance screening. Within each pipeline, daptomycin- and linezolid-resistant genomes (identified by AMRFinderPlus) were retained in full, with susceptible genomes randomly subsampled to match the resistant count (seed 88) to correct for sampling bias. These were combined with the LRE study isolates, the Queensland Genomics Health Alliance (QGHA) cohort (BioProject PRJNA797179), and E. faecium surveillance isolates from the Microbiological Diagnostic Unit Public Health Laboratory

(MDU PHL), Victoria. The final dataset of 1,880 genomes was submitted to PopPUNK for cluster assignment. DAP-R, daptomycin-resistant; LZD-R, linezolid-resistant; ANI, average nucleotide identity; ATB, AllTheBacteria.

A ≥99.8% ANI threshold was used to target the discrete intra-species evolutionary boundary (≥99.8% ANI) where strains share highly conserved functional gene content (“genomovars”) [27,28]. Because *E. faecium* exhibits a high baseline intra-species identity (mean ~99.03% [29]) and a highly plastic accessory genome (up to 38% [30]), this cutoff was necessary to restrict comparisons to near-clonal genetic backgrounds. This precision filter prevented the artificial merging of distinct clinical lineages, enabling high-resolution tracking of recent plasmid transmission without confounding noise from deep phylogenetic divergence.

#### S4. Characterisation of the *poxtA* genetic context

To investigate the genetic context of linezolid resistance, the *poxtA* gene and its flanking regions were extracted from the annotated assemblies. Initial screening involved extracting the *poxtA* coding sequence and 5,000 bp of upstream and downstream flanking sequences to identify conserved synteny. While the exact sequence matches the *E. faecium* reference allele *poxtA-Ef* in AMRFinderPlus (protein accession WP_094899500.1), it is functionally equivalent to the staphylococcal prototype [31] (see “Comparative Analysis with Tn6657, Tn6349, Tn554 and Tn6674” below for further details). For brevity, we use the family symbol *poxtA* throughout the text, though the database-specific *poxtA-Ef* label is retained in some figures and tables to maintain bioinformatic transparency.

To define the precise boundaries of the putative transposon, the terminal inverted repeats (IRs) of the flanking insertion sequences (IS) were determined using two distinct approaches. For the downstream IS*1678* element, reference IR sequences were retrieved from the ISfinder database [32]. For the upstream novel IS*1380*-family element, which was absent from public databases, we extracted 300 bp flanking sequence. These upstream and downstream flanking regions were aligned against each other using BLASTn (v2.15.0; [33]; with `-task` `blastn`) to identify the conserved inverted repeats based on sequence self-similarity. The identified IR sequences were then mapped to the *E. faecium* LRE_04 assembly using `seqkit locate` to define the full transposon coordinates [34].

The novelty of the identified transposon was assessed by searching the *poxtA*-carrying transposon sequence against the PLSDB plasmid database [35] and the ATB archive using LexicMap (v0.8.0; [36]). High-coverage (>60%) hits were retrieved, and their annotations were regenerated using Bakta to ensure consistent feature naming. In addition, we performed a targeted protein homology search using cblaster (v1.4.0; [37]). Briefly, cblaster queries the amino acid sequences of the transposon gene cluster against the NCBI non-redundant protein database to identify co-located homologous sequences. Hits were filtered to retain only those containing the complete transposon gene cluster.

Structural comparison of the transposon across the study isolates and external database hits was performed using Clinker (v0.0.32; [38]). Pairwise nucleotide identity between transposon variants was calculated using BLASTn (all-vs-all), with ANI weighted by alignment length. To detect direct target DNA repeats (DRs), the 12 bp immediately flanking the transposon boundaries were extracted and compared across all loci.

Tn*8026* is an 8,185 bp composite transposon with a GC content of 34.95%. It is flanked by two transposable elements of the IS*1380* family in direct orientation: IS*Efa26* (1,675 bp) at the 5' boundary and IS*1678* (1,320 bp) at the 3' boundary. The element is defined by Left and Right Inverted Repeats: IRL (26 bp: 5'-CCTGAATAATTCATAATTTTTCAAAA-3') and IRR (24 bp: 5'-AATTAAGCTATGAATATTCAGG-3'), which share a conserved 15 bp identical 5' terminal sequence (5'-CCTGAATAATTCATA-3'). An inventory of the 8 predicted ORFs of Tn8026 is presented in Table A1.

**Table A1: Predicted open reading frames (ORFs) of transposon Tn*8026*.**

| Locus Tag | Start | Stop | Strand | Gene | Product / Annotation |
|---|---|---|---|---|---|
| EKPHLE_01 | 155 | 1,468 | + | - | IS*1380* family transposase (IS*Efa26*) |
| EKPHLE_02 | 1,671 | 2,270 | - | *merR* | *MerR* family transcriptional regulator |
| EKPHLE_03 | 2,332 | 2,745 | + | - | GNAT family N-acetyltransferase |
| EKPHLE_04 | 2,789 | 3,310 | + | - | Transposase (accessory element) |
| EKPHLE_05 | 3,307 | 4,350 | + | - | IS*3* family transposase |
| EKPHLE_06 | 4,630 | 4,761 | + | - | Hypothetical protein |
| EKPHLE_07 | 4,851 | 6,485 | + | *poxtA-Ef* | ABC-F type ribosomal protection protein PoxtA-Ef |
| EKPHLE_08 | 6,663 | 7,982 | + | *tnp* | IS*1380* family IS*1678* transposase |

##### *Comparative Analysis with Tn6657, Tn6349, Tn554 and Tn6674*

Comparative sequence analysis shows that Tn*8026* represents a highly distinct lineage of *poxtA*-carrying transposons. BLASTn alignments between the backbones of Tn*8026* and Tn*6349* (*Staphylococcus aureus* clinical isolate; GenBank accession: MH746818.1; [39]) or Tn*6657* (the primary sub-module of Tn*6349* carrying *poxtA* and *fexB*) reveal no sequence similarity across their backbones (shared backbone coverage is 0%). Similarly, there is no sequence homology to the site-specific transposons Tn*554* and Tn*6674 [40,41]*. The comparative genomic differences are summarized in Table A2.

**Table A2 Genomic comparison on Tn*8026* against canonical reference elements**

| Transposon | Length (bp) | GC Content | Flanking Insertion Elements | Resistance Genes Carried | poxtA Sequence Identity (vs. Tn*8026*) |
|---|---|---|---|---|---|

| | | | | | |
|---|---|---|---|---|---|
| Tn*8026* | 8,185 | 34.95% | IS*Efa26* (5') / IS*1678* (3') (IS*1380* family) | *poxtA-Ef* | Reference (100%) |
| Tn*6657* | 14,435 | 36.25% | IS*1216* (flanking both ends) (IS*6* family) | *poxtA*, *fexB* | Nucleotide: 68.40% Protein: 74.07% |
| Tn*6349* | 48,350 | 35.45% | IS*1216*-like (flanking) (IS*6* family) | *poxtA, fexB, cfr, erm(B)* | Nucleotide: 68.40% Protein: 74.07% |
| Tn*554* | 6,691 | 32.60% | None (att554 insertion sites) (*tnpABC* recombinase) | *spc, ermA* | None (0% homology) |
| Tn*6674* | 12,932 | 33.89% | None (*radC* insertion sites) (*tnpABC* recombinase) | *optrA, fexA, spc, erm(A)* | None (1.34% *optrA*/*poxtA* domain homology) |

A significant divergence is identified in the *poxtA* gene itself. Tn*8026* carries the *poxtA-Ef* allele (1,635 bp), which exhibits only 68.40% nucleotide identity and 74.07% amino acid identity (536 amino acids aligned via BLASTp) with the canonical *poxtA* gene (1,632 bp) disseminated by Tn*6657* and Tn*6349* (these transposons are found in *S. aureus*). Furthermore, their accessory modules differ completely: Tn*6657* carries *fexB*, and Tn*6349* carries *cfr* and *erm(B)* inside its transposable boundary, whereas Tn*8026* carries a *MerR* transcriptional regulator, a GNAT family N-acetyltransferase, and two accessory transposases. This demonstrates that Tn*8026* represents a novel, structurally independent transposon lineage that has evolved to mobilise a highly divergent *poxtA* variant in *E. faecium*.

The *poxtA* allele carried by Tn*8026* corresponds to the *E. faecium* reference allele *poxtA-Ef* (WP_094899500.1; 99.4% amino-acid identity) and is notably divergent from the staphylococcal *poxtA* prototype carried by Tn*6657*/Tn*6349*, sharing only 72.8% amino-acid and 66.8% nucleotide identity (global, Needleman-Wunsch). This divergence reflects genuine allelic variation within the *poxtA* family rather than comparison to a paralogue, as confirmed by the markedly lower identity to *OptrA* (26.6%). Despite this divergence, the two *poxtA* alleles are functionally equivalent [31].

#### S5. Phylogenetic reconstruction and transmission analysis

Core genome clusters were defined using PopPUNK (v2.7.7; [42]) with the *E. faecium* v2 database. Transmission dynamics were investigated using the PopPIPE pipeline, which automates the analysis of bacterial population structure and transmission [43]. For high-resolution analysis of the study-specific clusters, we used TransPhylo (v1.4.5; [44]). TransPhylo reconstructs transmission trees from time-labeled phylogenies, allowing for the inference of both direct transmission events and those involving unsampled intermediate hosts. The probability of transmission between hosts and the number of unsampled intermediates (missing events) were inferred using a stochastic branching process model, incorporating collection dates derived from BioSample metadata.

Following the identification of the primary transmission cluster (Cluster 44), we performed high-resolution pairwise distance analysis using the Split K-mer Analysis (SKA) tool (v0.5.0; [45]). SKA was used to calculate alignment-free single nucleotide polymorphism (SNP) distances between all isolates in the cluster, which were visualised as a clustermap using the Seaborn (v0.13.2; [46]) library in Python.

#### S6. Linear plasmid comparative analysis

To investigate the relatedness and structural evolution of the novel linear plasmids identified in this study, linear plasmid sequences were extracted from the 24 LRE isolates and four Victorian (MDU) surveillance isolates (AUSMDU00056207, AUSMDU00067942, AUSMDU00074711, and AUSMDU00084267) identified as carrying similar linear replicons.

To resolve distinct structural variants, we performed a global synteny analysis using a custom MUMmer (v4.0.1; [47]) workflow. Pairwise alignments were generated for all plasmid pairs using `nucmer` with the `--maxmatch` parameter. To prevent repetitive elements, such as insertion sequences, from artificially inflating similarity scores, alignments were filtered to retain only 1-to-1 mappings using `delta-filter -1`.

We defined a "Global Identity" metric to penalise both sequence divergence and structural gaps as the total number of (1-to-1) matching bases, divided by the length of the longest sequence in the pair. Plasmids were clustered using Hierarchical Clustering (UPGMA/Average Linkage) based on a distance matrix derived from these identity scores (100 - global identity). Subcommunities were defined using a strict threshold of >99.5% global identity. A single representative was selected from each cluster (prioritising the longest sequence) for visualisation. Synteny plots were generated using pyGenomeViz (v1.6.1; [48]) with the `pgv-mummer` module.

We employed two additional approaches to validate the synteny-based clustering. First, we assessed structural rearrangement distances using Pling (v2.0.1; [49]) based on the Double Cut and Join Indel (DCJ-Indel) model, explicitly setting the topology to linear and defining subcommunities with a maximum distance threshold of 4 events. Second, to assess local nucleotide conservation independent of structural gaps, we calculated pairwise ANI using skani with a compression factor of 30 (`-c 30`) and a minimum aligned fraction of 60% for both sequences

To determine the global distribution and potential origin of the linear plasmid backbone, we screened our global dataset of *E. faecium* genomes using LexicMap. The modal representative of the study cohort, pELF_LRE_29 (Cluster 3; 132,853 bp), was selected as the query sequence to ensure search results reflected the dominant circulating genotype. The query was searched against an index of the global phylogeny dataset. To identify high-confidence homologues while allowing for structural variation, we applied a minimum query coverage threshold of 70% per genome and a minimum alignment length of 2000 bp. In addition, we increased the seed minimum prefix lengths (`-p 19 -P 21`) to maintain sensitivity for high-similarity subjects while improving search performance.

Lastly, to determine whether the entire linear plasmid had transferred across species boundaries, the short-read assemblies of the two Tn*8026*-positive *E. gallinarum* isolates from South Korea (GCF_030340145.1 and GCF_030340105.1) were aligned to the dominant outbreak reference plasmid, pELF_LRE_29, using minimap2 with the `asm5` preset. Total linear plasmid coverage was subsequently quantified by merging overlapping alignment intervals.

##### *S6.1 Characterising linear plasmids as pELF-like*

To substantiate the taxonomic classification of the cohort linear plasmids as enterococcal linear form-like (pELF-like) vectors, we performed comparative sequence analysis of our modal representative plasmid, pELF_LRE_29 (132,853 bp), against the canonical reference family members pELF1 (143,316 bp; GenBank: LC495616.1; [50]) and pELF2 (108,102 bp; GenBank: AP022343.1; [51]).

When compared via BLASTn alignment, pELF_LRE_29 showed extensive structural conservation with pELF1, sharing 90,487 bp of sequence at 99.36% weighted nucleotide identity (68.11% coverage on pELF_LRE_29 and 63.14% on pELF1). This overlap exceeds the backbone conservation observed in recently described global pELF1-like variants, which typically share >75 kb of core architecture at >97% identity with pELF1 [52]. Alignment against the smaller pELF2 reference revealed a shared sequence length of 63,552 bp at 98.51% weighted identity, spanning 58.79% of the pELF2 sequence (47.84% coverage on pELF_LRE_29). In both comparisons, the unaligned regions correspond to mobile accessory insertions—predominantly transposon-mediated antimicrobial resistance determinants—embedded within a highly syntenic, conserved core pELF backbone.

This classification is further supported by a genomic signature of the pELF family: a distinctly lower base composition relative to the host chromosome. The pELF_LRE_29 contig has a GC content of 34.51%, representing a 3.38% decrease compared to the chromosome (37.89% GC). This closely matches the established global family mean of ~33.9% defined by Hashimoto et al. (2023) [53].

Functional annotation of the pELF_LRE_29 sequence confirmed the strict preservation of the universally conserved pELF core pangenome required for linear plasmid replication, partitioning, vertical stability, and horizontal transfer [52,53]. Replication initiation is driven by multiple *repB* genes belonging to the characteristic Rep_2 and Rep_3 superfamilies, including an initiator Rep protein containing a WH1 domain (located at coordinates 9,552–10,319 bp, 10,355–11,515 bp, and 12,564–13,367 bp) [54]. Active partitioning and segregation machinery are encoded by a Type II active system comprising a *ParM*-like actin ATPase domain (1,765–2,820 bp) and a *Soj/ParA*-like AAA domain-containing ATPase (3,675–4,493 bp) [52]. Consistent with a linear topology requiring coordinated replicon segregation during cellular division, an *ftsK* DNA translocase domain-containing protein was identified at coordinates 92,860–94,236 bp [55].

Vertical inheritance is reinforced by a high density of Type II Toxin-Antitoxin (TA) modules distributed across the backbone, including *relB-relE* (4,719–5,208 bp), multiple *hicA-hicB* systems (7,247–7,894 bp and 8,963–9,361 bp), an epsilon-*parD* complex (64,803–66,861 bp), and a *relB*/*dinJ-mazF* module (131,774–132,386 bp) [53]. Horizontal transfer capability is mediated by a highly conserved conjugative *tra* operon spanning approximately 110 kb

(22,166–132,386 bp, with accessory resistance insertions masked), encoding 39 transfer-associated genes [56].

#### S7. Structural Analysis of the *vanA* Gene Cluster (Tn*1546*)

Genetic characterisation of the *vanA* resistance region revealed two primary configurations of the Tn*1546* transposon, designated here as Group I and Group II (Figure S3). Both configurations diverged from the prototypical Tn*1546* structure [57] (GenBank accession M97297) through the incorporation of multiple insertion sequences, specifically IS*1542* and IS*1521*.

The predominant configuration (Group I), identified in most isolates (24/28; primarily ST80), featured a 5' upstream region containing an IS*1216E* element, an rRNA adenine N-6-methyltransferase, and the macrolide resistance gene *erm(B)*. This was followed by a transposase sharing 81% identity with the Tn*1546* ORF1 (Tn*3* family transposase). The core *van* cluster in this group was characterised by the insertion of an ISL*3*-family IS*1521* transposase between *vanS* and *vanH*, and an IS*256*-family IS*1542* transposase between the resolvase (ORF2) and *vanR*. The 3' region was conserved across Group I isolates, encoding an HTH ArsR-type transcriptional regulator, a SpoVT-AbrB domain protein, and the ParE toxin of the ParDE type II toxin-antitoxin system. Minor structural variations were observed within this group: LRE_16 harboured a truncated *vanS* gene (36% identity to reference), while isolates LRE_08 and LRE_10 contained additional hypothetical or LPXTG cell wall anchor domain proteins immediately upstream of the 5' IS*1216E* element.

The second configuration (Group II) was exclusive to the ST1424 lineage (LRE_05, LRE_09, and LRE_20) and displayed significant structural degradation, including the loss of the initial transposase, resolvase, and the accessory *vanZ* gene. The upstream region in Group II was defined by an IS*1216E* element followed by an IS*256*-family transposase (85% similarity to

IS*1542*). Internally, these isolates harboured a second IS*1216E* element inserted in the reverse orientation between *vanX* and *vanY*, followed by a forward-strand transposase downstream of *vanY*. Unlike Group I, the downstream region contained a cation:proton antiporter rather than the ParDE system. Notably, isolate LRE_05 exhibited further plasticity with the complete loss of *vanY* from the van cluster; genomic analysis located *vanY* on a separate plasmid immediately flanked by an IS*1216E* transposase, suggesting a recent, distinct mobilisation event followed by the acquisition of a putative replication protein downstream of the main cluster. Given the extensive deletion of the transposition machinery and the diverse upstream/downstream contexts of Group II compared to Group I, this region likely represents a distinct genetic element or a remnant of a separate transposition event rather than a direct derivative of the Tn*1546* transposon found in the Group I isolates.

#### S8. Discordant linezolid phenotype-genotype results

Etest validation revealed a divergence in these phenotypes: LRE_05 displayed a borderline MIC of 4 mg/L, whereas LRE_20 exhibited high-level resistance with an MIC of 16 mg/L. The profile of LRE_05 mirrors findings from recent surveillance studies, where isolates initially classified as resistant by automated systems were found to be susceptible (MIC ≤ 4 mg/L) upon re-testing, suggesting that non-standardised AST methods contribute substantially to genotype-phenotype disagreement in the absence of acquired genes [58]. However, the high-level resistance confirmed in LRE_20 cannot be attributed to method instability and remains unexplained by current databases, pointing to the existence of a novel or uncharacterised resistance mechanism within this lineage.

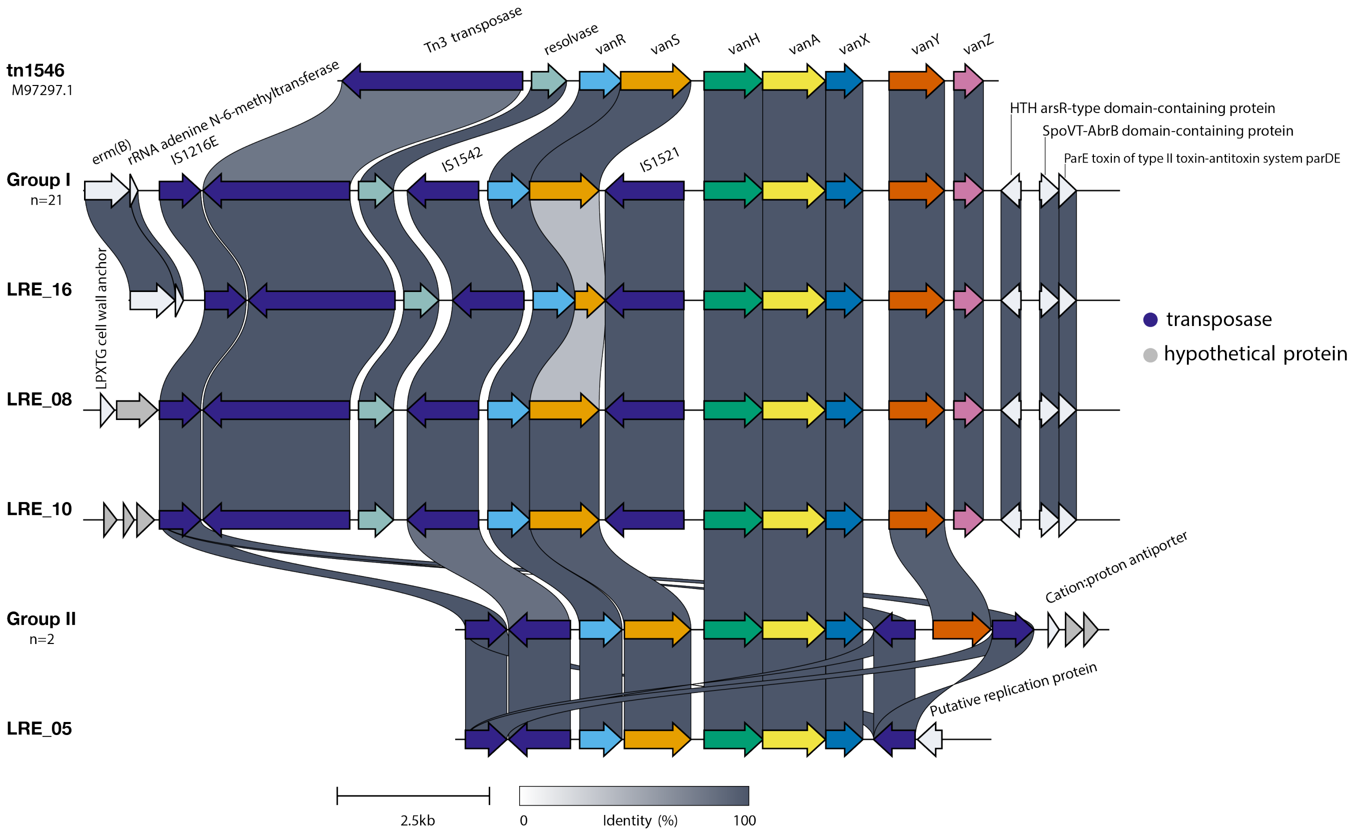


**Figure S3: Structural comparison of the *vanA* gene cluster and flanking regions across *E. faecium* study isolates.** Linear comparison of the *vanA* resistance region in representative Group I (ST80) and Group II (ST1424) isolates, alongside variants LRE_16 and LRE_05, aligned against the prototypical Tn*1546* transposon (GenBank accession M97297). Arrows represent open reading frames (ORFs) indicating the direction of transcription and are coloured by functional category. Grey shaded links between tracks indicate identity. Key structural features highlighted include the insertion of IS*1521* and IS*1542* within the *van* cluster of Group I, the truncation of *vanS* in LRE_16, the extensive 5' deletion and loss of *vanZ* in the ST1424 lineage (Group II), and the distinct excision of the *vanY* gene in isolate LRE_05.

#### S9. Detailed reconstruction of Tn*8026* intercontinental and interstate transmission dynamics

To infer the transmission history of this cluster, we generated a consensus transmission tree using TransPhylo (Figure 3; [44]). The model estimated the date of the index case for Cluster 44 to be circa 2014. This estimated emergence postdates the earliest global detection of Tn*8026* in a historical isolate from Norway (2012), suggesting that the transposon was already established in the global *Enterococcus* population prior to the expansion of the specific outbreak lineage described here.

Transmission analysis within Cluster 44 revealed a high-probability linkage connecting isolates from India, Victoria, and the Queensland LRE cohort. Specifically, TransPhylo analysis inferred a directional transmission event originating from the Indian isolate SAMN32671899 (Puducherry, 2021) to the Victorian isolate SAMN45173172 (AUSMDU00068490; 01/2022) with a posterior probability of 1.0 (Figure 3, Figure S4). This epidemiological link was supported by high-resolution genomic comparison, which revealed that the two isolates differed by only 11 SNPs, confirming they are effectively the same clone (Figure S5). Notably, both isolates harbour the novel transposon Tn*8026* with 100% coverage, providing definitive genomic support for the epidemiological link inferred by the model.

Crucially, this computational inference is independently corroborated by epidemiological records. Patient data associated with the Victorian cohort revealed documented travel histories to India [22], supporting a scenario of repeated plasmid importation rather than purely local clonal expansion. The transmission chain subsequently extended to SAMN45173170 (AUSMDU00067942; 01/2022), a Victorian isolate previously characterised in this study (Group II, Figure 1) as carrying Tn*8026* on a linear plasmid. These two Victorian isolates were separated by only 2 SNPs (Figure S5). Collectively, these genomic and epidemiological data confirm that the Tn*8026*-carrying linear plasmid vector was imported from the Indian subcontinent and subsequently established local circulation in Victoria.

Furthermore, the transmission reconstruction linked this international/interstate chain to the Queensland LRE cohort. The analysis inferred a linkage between the Indian isolate

SAMN32671899 and the Queensland isolate LRE_23. However, this link was not direct; the model inferred the presence of two unsampled intermediate hosts separating SAMN32671899 from LRE_23, with a genetic distance of 29 SNPs between the two (Figure S5). This "missing link" topology is consistent with the introduction of the strain into Queensland via unmonitored routes or asymptomatic carriage prior to its detection in the LRE outbreak.

#### S10. Synteny analysis of the linear plasmid

Structural diversification within the cohort was driven by presumed sequential deletion events occurring at the "left" or hairpin end of the plasmid, immediately upstream of the Tn*1546* transposon. Cluster descriptions can be found in Table S7. The first major variation was observed in Cluster 7 (pELF_LRE_10 and pELF_LRE_02). While sharing 100% internal identity, these ~120 kb plasmids are distinguished from the core lineage by the loss of a ~10.6 kb MDR region located immediately upstream of Tn*1546*. This deleted module contains the resistance genes *cfr*(D) (linezolid/phenicol resistance) and *erm(B)* (macrolide resistance), along with Apt and GMP synthase. The region is flanked by IS*1216E* transposases, forming a structure characteristic of a composite transposon. This architecture suggests the deletion may have resulted from the excision of a novel, uncharacterised mobile element, or via homologous recombination between the flanking IS copies.

A further reductive evolution event was identified in the singleton Cluster 8 (pELF_LRE_08). This ~108 kb plasmid appears to be a minimal variant of the lineage. In addition to lacking the *cfr*(D)/*erm(B)* module, it has lost a second ~13.3 kb cargo region located immediately upstream of the MDR deletion site (towards the hairpin end). This deletion removes a region containing an MFS transporter, a phage protein, an XRE-family transcriptional regulator, and multiple hypothetical proteins.

The remaining two Victorian isolates formed distant singleton clusters with significantly divergent resistance region architectures, sharing <85% global identity with the LRE cohort. Cluster 10 (AUSMDU00056207) carries a truncated form of Tn*1546*, lacking the initial Tn*3*-family transposase and resolvase genes and missing Tn*8026* entirely. In contrast, Cluster 9 (AUSMDU00084267) lacks Tn*1546* entirely but retains a copy of Tn*8026*, albeit located towards the hairpin end rather than the open end as observed in the main cohort. However, this Tn*8026* element is structurally distinct from that found in the other clusters, containing an IS*1251* insertion between the *poxtA* gene and the right boundary IS*1678*, consistent with the variant structure identified earlier (Figure 4).

##### *Complementary structural validation*

These findings were supported by the DCJ-Indel analysis using Pling, which similarly identified the structural divergence of pELF_LRE_02 (Cluster 7) from the main LRE group (distance of 5–7 events; Figure S7). However, pairwise ANI analysis using skani revealed that despite these large structural deletions, the remaining backbones share exceptionally high nucleotide identity (>99.9%) across the entire cohort (Figure S8). This confirms that the observed structural diversity is not due to the acquisition of unrelated plasmids, but rather the reductive evolution of a single, highly conserved linear plasmid backbone.

##### *Global reservoir and importation of the linear plasmid*

To determine the global distribution and potential evolutionary origins of this conserved linear backbone, we screened the global dataset used for the phylogeny with LexicMap, as well as PLSDB. The screen identified the linear plasmid backbone in 45 unique genomes from the global dataset (Table S6), but none from PLSDB. After excluding the 28 isolates from the Australian study cohort (24 LRE from this study and 4 Victorian), 17 external genomes were identified with the plasmid. These hits revealed a widespread distribution, with the backbone detected in isolates from Australia (historical Queensland, *n*=10), China (*n*=3), India (*n*=2), Bangladesh (*n*=1), and Norway (*n*=1).

Structural analysis of these global hits supports the hypothesis that the multidrug-resistant LRE plasmid emerged from a broadly circulating linear backbone. While the complete pELF_LRE_29 architecture (>90% coverage) was unique to the Australian outbreak cohort, historical Australian isolates (2019) and Chinese isolates (2021–2022) carried conserved variants sharing 70–79% of the query sequence. Notably, the detection of the backbone in ten historical Queensland isolates from 2019 indicates that variants of this linear plasmid have been circulating locally in a non-linezolid resistance form for at least four years prior to the current outbreak.

Despite this established local reservoir, the genomic data strongly supports an independent importation event for the specific lineage driving the current outbreak. The LexicMap screen identified the Indian isolate SAMN32671899 (Puducherry, India, 2021) as carrying a distinct, high-coverage variant of the linear plasmid (81.9% query coverage)— higher than the historical Queensland background (72–78%)—including Tn*8026*. This finding aligns perfectly with the consensus transmission tree (Figure 3), which independently identified SAMN32671899 as the direct ancestor of the Australian cluster.

The plasmid coverage data mirrors the transmission reconstruction, showing a stepwise acquisition of content along the importation route: from the Indian progenitor SAMN32671899 (81.9%) to the intermediate Victorian host SAMN45173172 (80.7%), and finally to the subsequent Victorian isolate SAMN45173170 (93.8%), which clustered firmly within the LRE structural group (Cluster 6). This progression suggests that while the linear backbone is endemic to Queensland, the specific linezolid-resistant outbreak strain was likely imported into Victoria from the Indian subcontinent as a distinct "pre-LRE" lineage. It subsequently acquired additional structural modifications—increasing its similarity to the final LRE type—before its eventual transmission to Queensland and detection in isolate LRE_23. However, we note that without closed, long-read plasmid sequences from the historical and international short-read datasets, the precise structural events driving this evolution cannot be fully resolved.

The global synteny analysis partitioned the 28 linear plasmids into 10 clusters, revealing a dominant ~132 kb lineage and distinct deletion variants. Most isolates (*n*=20) belonged to a highly conserved "Core LRE Complex" (Clusters 1–6) characterised by a shared ~132 kb backbone carrying a full complement of resistance and cargo genes. This core complex includes the reference plasmid pELF_LRE_17 (Cluster 1) and two major sub-lineages represented by pELF_LRE_29 (Cluster 3) and pELF_LRE_15 (Cluster 6), which share ~99.0% global identity. Notably, the Victorian surveillance isolates AUSMDU00074711 and AUSMDU00067942 clustered firmly within this complex (Clusters 3 and 6, respectively), indistinguishable from the primary Queensland outbreak plasmids.

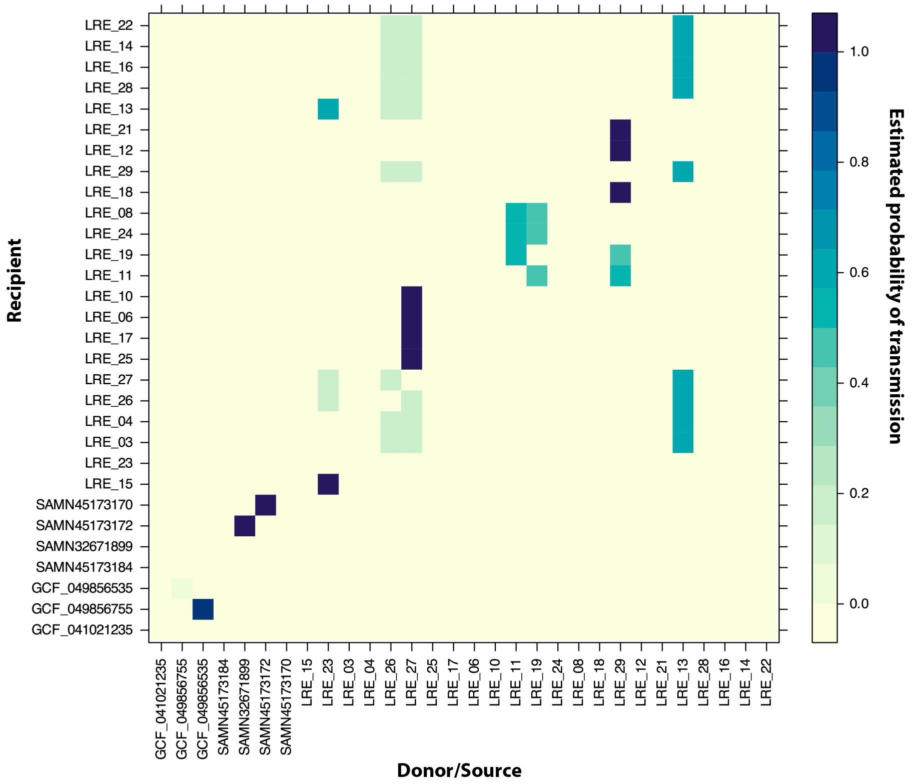


**Figure S4. Pairwise transmission probability matrix for the *poxtA*-positive *E. faecium* Cluster 44.** Heatmap displaying the posterior probabilities of direct transmission between sampled isolates as inferred by TransPhylo. The x-axis represents the potential donor (source) isolate, and the y-axis represents the potential recipient isolate. The colour intensity of each cell corresponds to the estimated probability of transmission from the sample on the x-axis to the sample on the y-axis, ranging from 0 (lowest probability) to 1 (highest probability).

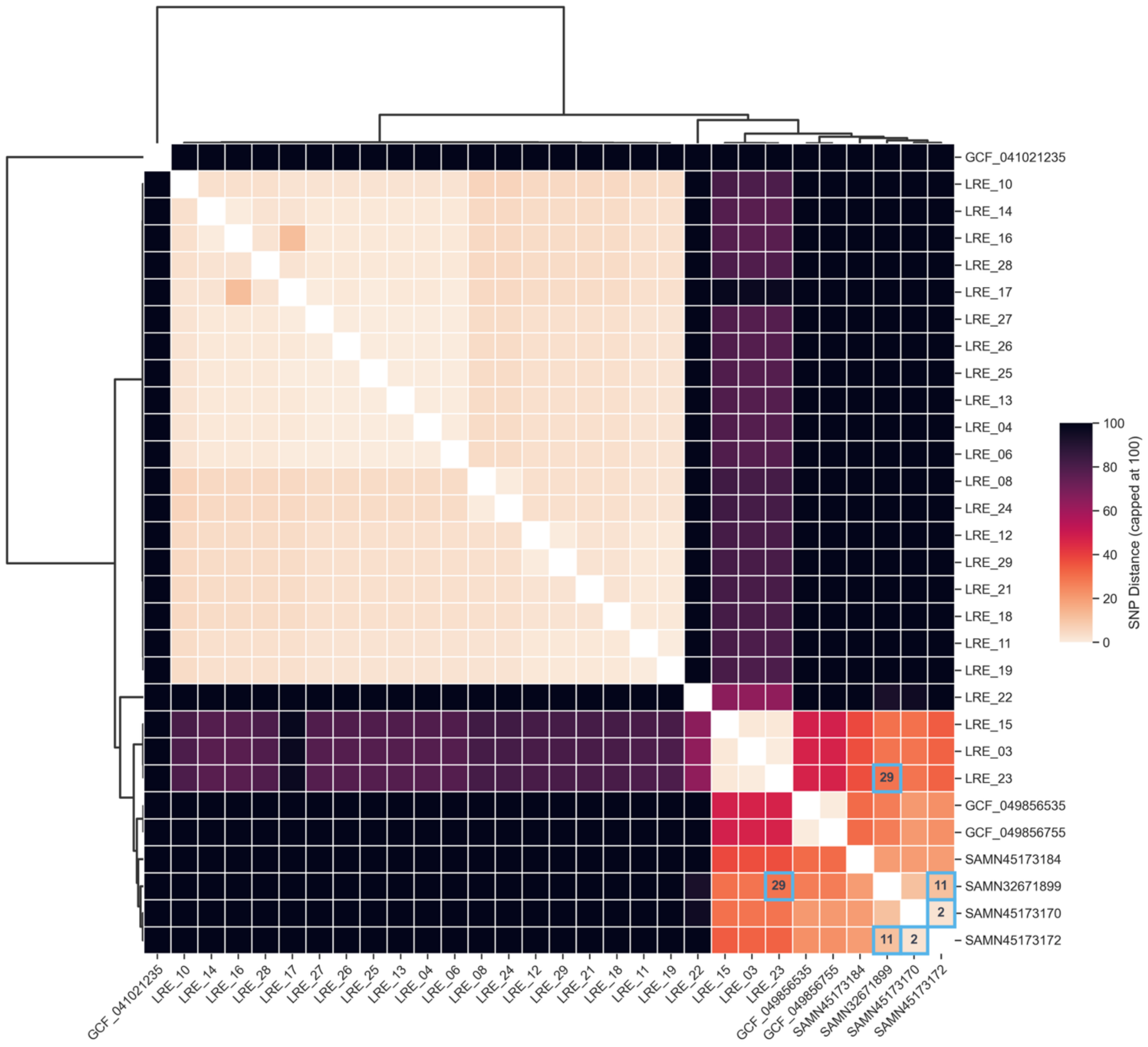


**Figure S5. Clustermap of pairwise SNP distances within the *poxtA*-positive *E. faecium* Cluster 44.** Clustermap displaying the pairwise SNP distances between isolates, calculated using SKA. The colour scale represents the genetic distance, capped at 100 SNPs to highlight fine-scale differences between closely related strains. Hierarchical clustering (average linkage) groups isolates by genetic similarity. Coloured boxes highlight key transmission pairs identified in the consensus transmission tree (Figure 2): **(A)** The transmission from India (SAMN32671899) to Victoria (SAMN45173172), separated by 11 SNPs; **(B)** The local spread within Victoria (SAMN45173172 to SAMN45173170), separated by 2 SNPs; and **(C)** The link between the Indian lineage and the Queensland LRE cohort (SAMN32671899 to LRE_23), separated by 29 SNPs. Values inside the cells indicate the exact SNP count.

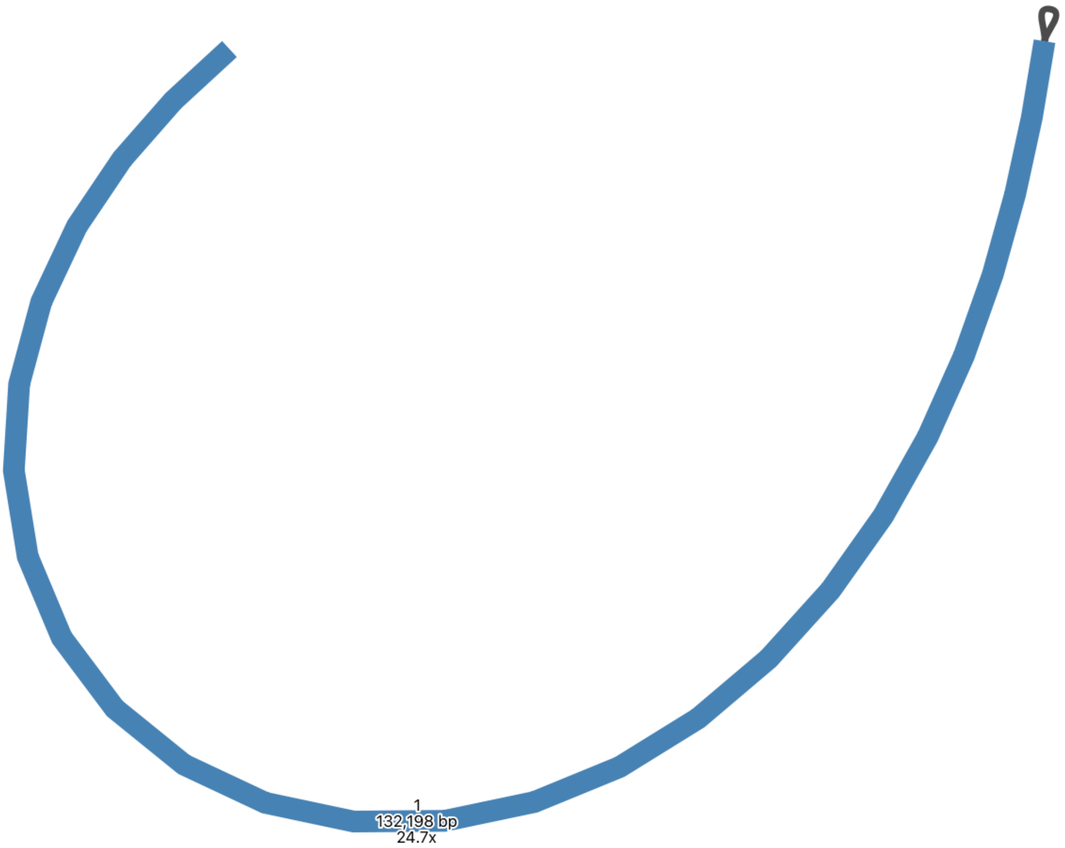


**Figure S6. Hybrid termini of linear plasmid.** A view of the assembly graph for the linear plasmid of sample LRE_29. This layout of the termini is consistent with all linear plasmids in this study: a hairpin end, where the terminus folds back on itself, and a protein-capped (open) end.

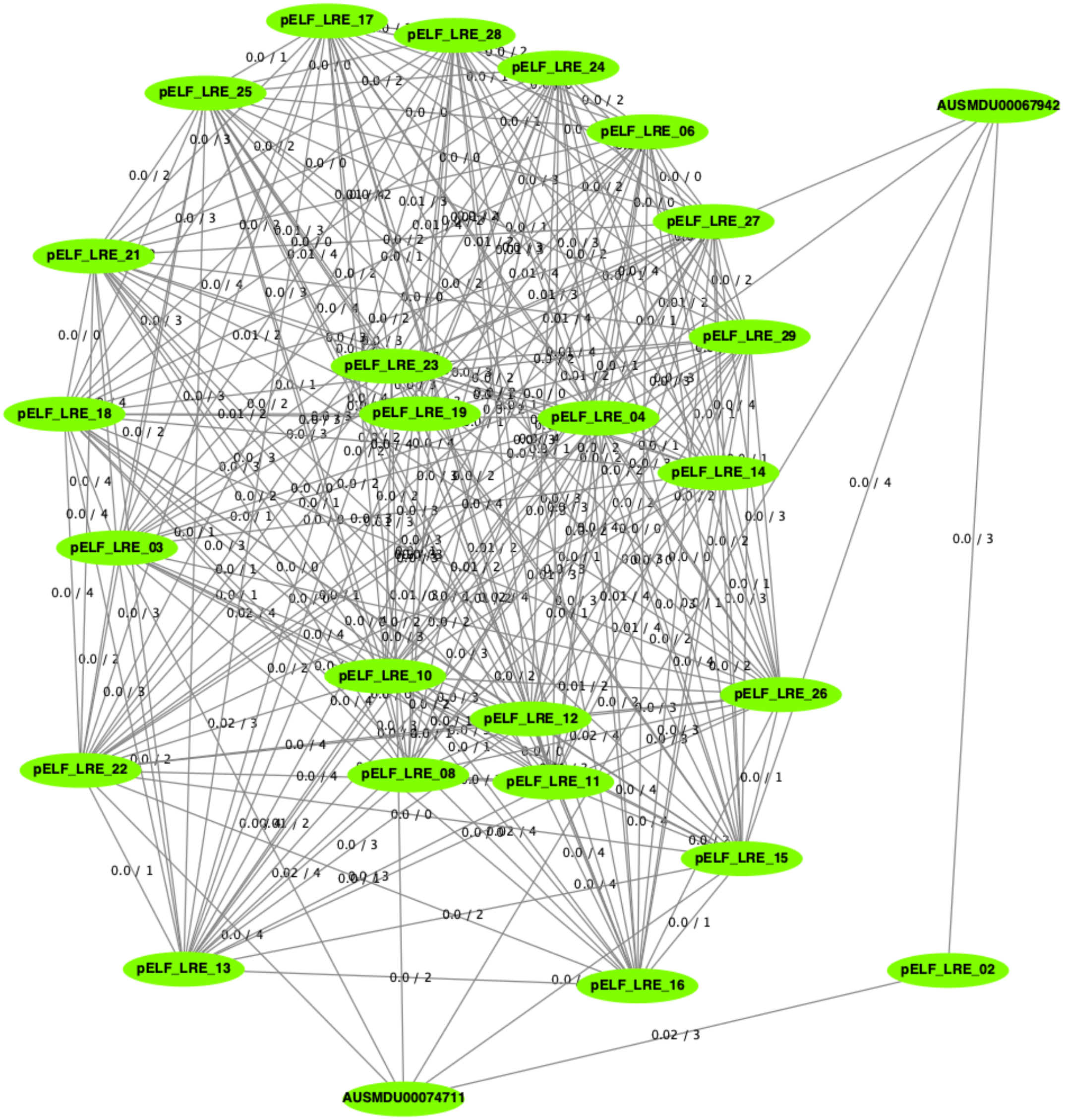


**Figure S7. Pling rearrangement distance network for linear plasmids.** Green nodes represent the linear plasmid of a sample, with the edges showing the containment and rearrangement distances (containment/rearrangement). No edge indicates the rearrangement distance was >4.

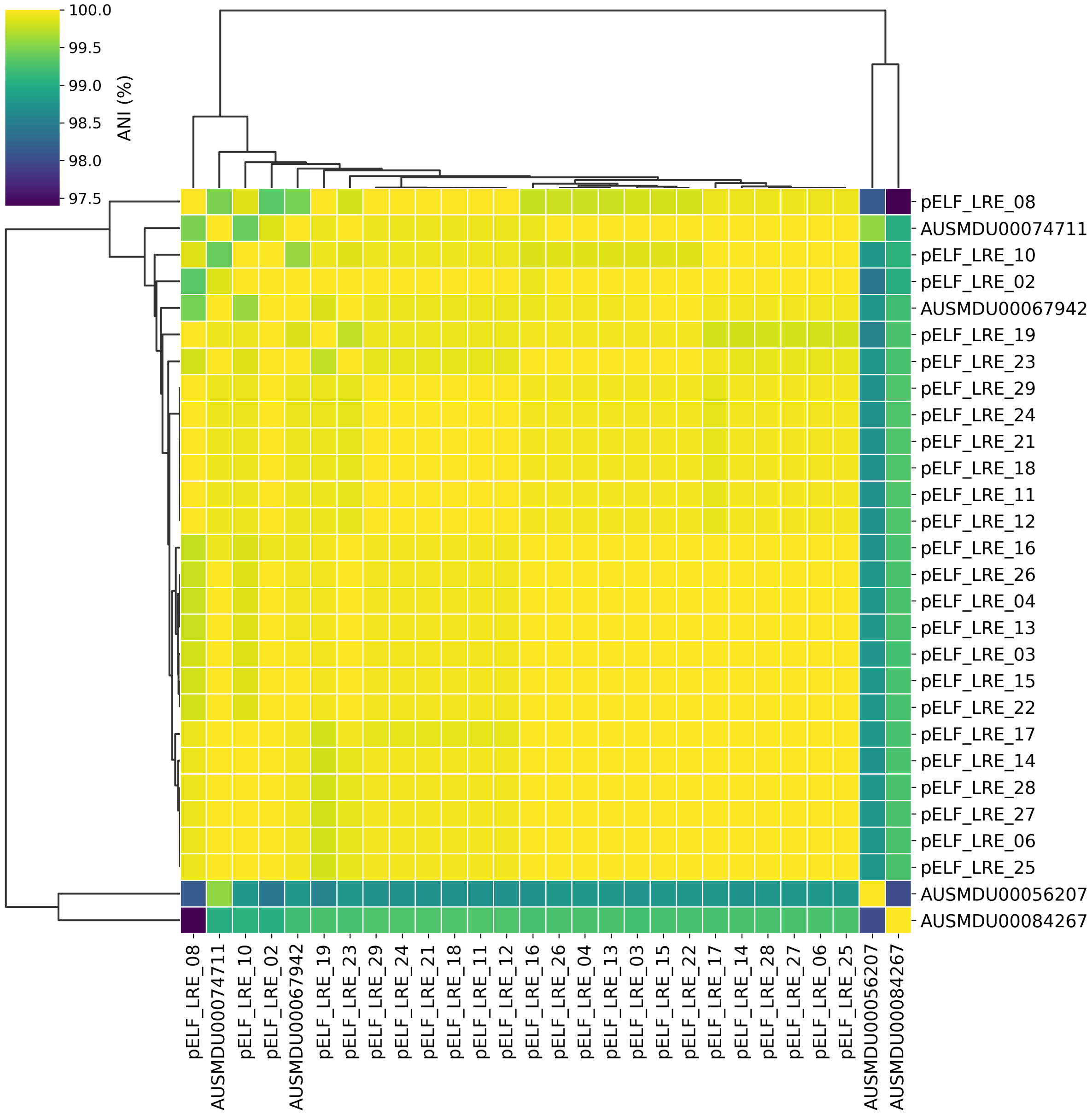


**Figure S8. Pairwise Average Nucleotide Identity (ANI) of linear plasmids.** Heatmap displaying the pairwise ANI between linear plasmid sequences extracted from the LRE cohort (*n*=24) and Victorian surveillance isolates (*n*=4). ANI values were calculated using skani and clustered using hierarchical clustering. The colour scale represents the percentage identity, ranging from blue (lower identity) to red (higher identity).

2. Hasman H, Clausen PTLC, Kaya H, Hansen F, Knudsen JD, Wang M, et al. LRE-Finder, a Web tool for detection of the 23S rRNA mutations and the optrA, cfr, cfr(B)

and poxtA genes encoding linezolid resistance in enterococci from whole-genome sequences. J Antimicrob Chemother. 2019;74: 1473–1476. doi:10.1093/jac/dkz092

54. Almeida-Santos AC, Tedim AP, Duarte B, Silva LM, Teixeira J, Castro AP, et al. Unnoticed spread of linear VanA-plasmids in vancomycin-variable Enterococcus

faecium strains across different regions: a diagnostics challenge. J Antimicrob Chemother. 2025; dkaf409. doi:10.1093/jac/dkaf409

55. Boumasmoud M, Dengler Haunreiter V, Schweizer TA, Meyer L, Chakrakodi B, Schreiber PW, et al. Genomic Surveillance of Vancomycin-Resistant Enterococcus faecium Reveals Spread of a Linear Plasmid Conferring a Nutrient Utilization Advantage. mBio. 2022;13: e03771-21. doi:10.1128/mbio.03771-21

56. Kurushima J, Ota N, Yoshii Y, Tomie N, Tomita H. A Novel Conjugation System in AMR-associated pELF-type Linear Plasmids of Enterococcus faecium. bioRxiv; 2026. p. 2026.01.27.701956. doi:10.64898/2026.01.27.701956

57. Arthur M, Molinas C, Depardieu F, Courvalin P. Characterization of Tn1546, a Tn3-related transposon conferring glycopeptide resistance by synthesis of depsipeptide peptidoglycan precursors in Enterococcus faecium BM4147. J Bacteriol. 1993;175: 117–127. doi:10.1128/jb.175.1.117-127.1993

58. Coll F, Gouliouris T, Blane B, Yeats CA, Raven KE, Ludden C, et al. Antibiotic resistance determination using Enterococcus faecium whole-genome sequences: a diagnostic accuracy study using genotypic and phenotypic data. Lancet Microbe. 2024;5: e151–e163. doi:10.1016/S2666-5247(23)00297-5
